# Distinct bacterial hosts, shared *bla_OXA-48_* plasmid backbones: longitudinal comparative genomics of carbapenemase-producing Enterobacterales from hospital wastewater, biofilms and patients

**DOI:** 10.64898/2026.08.15.26360502

**Authors:** Morgane Roger-Margueritat, Verena Schmidt, Gregory E. McCallum, Eva Gendron, Patrice Morand, Claire Terreaux-Masson, Caroline Landelle, James P. J. Hall, Aurélie Hennebique, Elena Buelow

## Abstract

Hospital wastewater (WW) and wastewater biofilms (WWB) are increasingly recognized as important reservoirs of carbapenemase-producing Enterobacterales (CPE), yet their long-term ecological dynamics and relationship with contemporaneous clinical isolates remain poorly understood. Here, we performed longitudinal CPE surveillance of WW and WWB over a 17-month period, combining culture-based screening and comparative whole-genome sequencing of environmental isolates with CPE isolates recovered from patients hospitalized in the same hospital building. A total of 42 environmental and 21 clinical CPE isolates were characterized. Environmental CPE populations underwent a marked ecological shift, with *bla_OXA-48_*-producing *Citrobacter* spp. progressively replaced by *bla_VIM-4_*-producing *Serratia nevei*. In contrast, clinical isolates remained taxonomically diverse throughout the study period, with a range of betalactamases including *bla_OXA-48_*, *bla_VIM-4_*, and *bla_NDM_*, with no comparable temporal replacement. Comparative genomic analyses revealed a strong association between resistance genes and mobile genetic elements (MGEs), with MGE dynamics largely following those of their hosts. *bla_OXA-48_* was predominantly associated with highly conserved IncL/M plasmid backbones shared across environmental and clinical compartments, whereas *bla_VIM-4_* was consistently embedded within conserved class 1 integron-associated genetic contexts on IncHI2A-rep1088 plasmids. In contrast, *bla_NDM_* displayed heterogeneous genomic organizations involving multiple plasmid backgrounds and frequent chromosomal integration. Together, our findings show that bacterial hosts and carbapenemase-carrying genetic elements follow distinct ecological trajectories within hospital WW ecosystems. Integrating longitudinal environmental surveillance with comparative genomics provides new insights into the persistence of clinically important carbapenemases across interconnected environmental and clinical reservoirs.

**Graphical Abstract:** 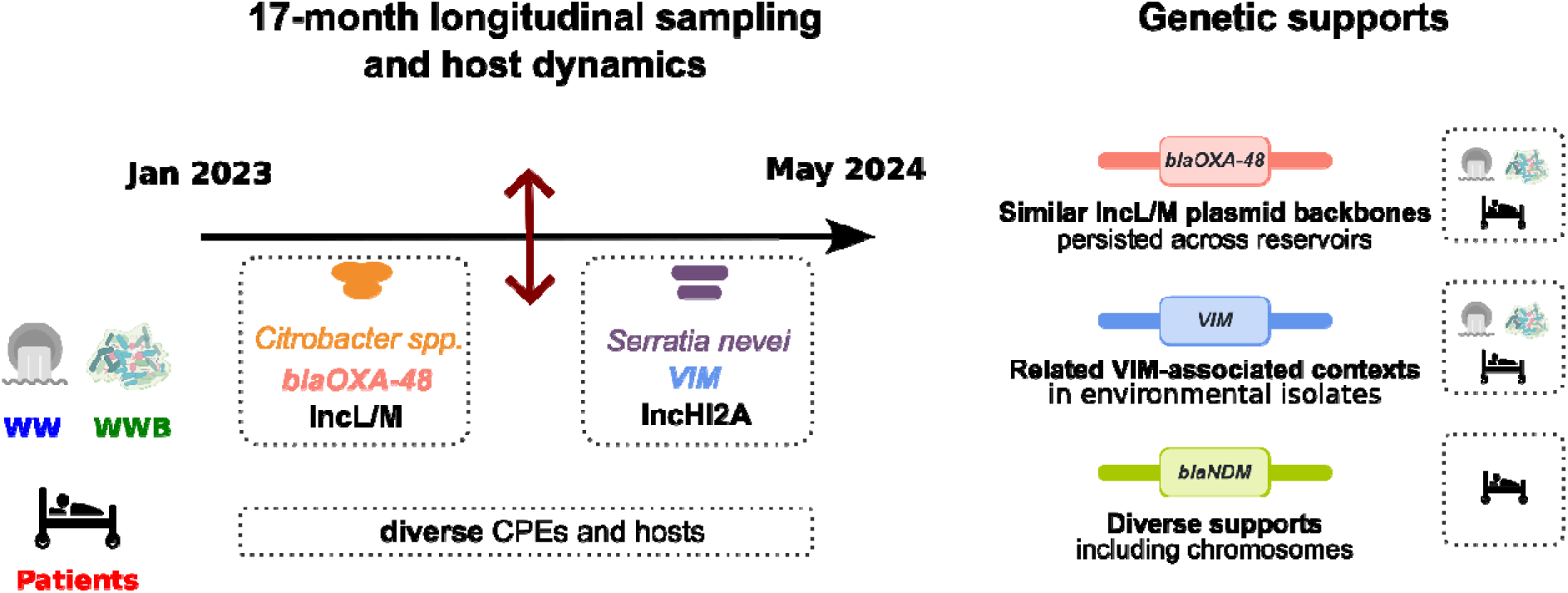

## Introduction

Hospital wastewater (WW) systems are increasingly recognized as important reservoirs of antimicrobial resistance (AMR), continuously receiving antibiotic residues, resistant bacteria and human-associated microorganisms from healthcare activities (Anantharajah et al., 2024; Hennebique et al., 2025). Within these systems, WW biofilms (WWB) constitute stable multispecies communities that can promote bacterial persistence, interspecies interactions and horizontal gene transfer (HGT), thereby potentially contributing to the long-term maintenance and dissemination of antibiotic resistance genes (ARGs) (Abe et al., 2020; Buelow et al., 2023). Among resistant pathogens, carbapenemase-producing Enterobacterales (CPE) are of particular concern due to their resistance to carbapenems, which are often considered last-resort antibiotics for severe Gram-negative infections (Logan and Weinstein, 2017). Carbapenemase genes, including *bla_NDM_*, *bla_OXA-48_*, *bla_VIM-4_*, *bla_KPC_* and *bla_IMP_*, are frequently associated with mobile genetic elements (MGEs), such as plasmids and transposons, facilitating their dissemination across bacterial species and environmental compartments (Mathers et al., 2011; Partridge et al., 2018).

Although hospital WW -associated biofilms are increasingly investigated as environmental AMR reservoirs (Buelow et al., 2023; De Geyter et al., 2017; Hennebique et al., 2025), important knowledge gaps remain regarding the temporal dynamics and ecological persistence of CPE populations within these systems. Most studies have focused on short-term surveillance or bulk resistome characterization, while the long-term evolution, replacement and persistence of carbapenemase-producing populations in WW ecosystems remain insufficiently understood. In particular, the potential role of WWB as long-term reservoirs supporting the maintenance and circulation of plasmid-associated carbapenemase genes has received limited attention.

Characterizing the genetic context of carbapenemase dissemination in environmental reservoirs also remains challenging (Mathers et al., 2024). Carbapenemase genes are frequently carried by plasmids and other MGEs with complex and dynamic architectures that are difficult to reconstruct using conventional short-read sequencing approaches (M et al., 2025). Recent advances in whole genome sequencing (WGS), including long-read sequencing technologies, now provide improved opportunities to characterize ARG-associated plasmids, investigate resistance dissemination and explore evolutionary relationships between environmental and clinical bacterial populations (Arredondo-Alonso et al., 2017; Bortolaia et al., 2020; Ludden et al., 2017). In this context, the present study investigated the temporal dynamics of CPE populations in hospital WW and WWBs from Grenoble Alpes University Hospital over a 17-month period. Using culture-based isolation, antimicrobial susceptibility testing and WGS, we characterized the evolution of CPE populations and associated resistance determinants in environmental compartments and compared them with patient-derived CPE isolates collected during the same period. This work provides new insights into the ecological dynamics and persistence of carbapenemase-producing populations within hospital WW ecosystems.

## Materials and Methods

### 2.1 Study setting

The study was conducted at Grenoble Alpes University Hospital (CHUGA), France, the reference university hospital of the Isère department (South-East region of France). CHUGA comprises 2,298 hospital beds and serves a population of approximately 675,000 inhabitants. The ICU building includes a 20-bed medical Adult Intensive Care Unit (AICU), a 15-bed surgical AICU, a 12-bed neurosurgical AICU, a 10-bed medical Adult Intermediate Care Unit (AIMCU), and a 12-bed surgical AIMCU. These units are located within the same building and share a common WW collection system from which the WW and WWB samples of this study have been collected. All AICU rooms are single occupancy.

### 2.2 Environmental sampling

Between January 2023 and May 2024, a total of 23 WW and WWB samples were collected from the WW outlet of the AICU at the CHUGA. Among the 23 samples, 14 were WW and 9 were WWB samples. WW samples were collected by inserting open 50 mL sterile centrifuge tubes (Corning) attached to a string into the WW pipelines. WWB samples were obtained by inserting closed 50 mL sterile centrifuge tubes (Corning) into the same pipelines and leaving them in place for one month to allow biofilm formation on the tube surface. Following retrieval, biofilms were collected by scraping the outer surface with a sterile cell scraper and subsequently resuspended in 1× phosphate-buffered saline (PBS; 137 mM NaCl, 2.7 mM KCl, 10 mM Na₂HPO₄, 1.8 mM KH₂PO₄, pH 7.4). All collected samples were transferred to 50 mL high-performance centrifuge tubes (VWR®) and centrifuged at 7,000 rpm for 15 min. Supernatants were discarded and pellets were resuspended in 5 mL PBS. A 1 mL aliquot of each suspension was mixed with glycerol at a final concentration of 20% and stored at −80°C until further analysis.

### 2.3 Culture-based isolation and characterization of environmental bacterial isolates

#### 2.3.1 Isolation of aerobic flora and carbapenemase-producing organisms

To assess the diversity of cultivable aerobic bacteria and screen for clinically relevant multidrug-resistant organisms, both non-selective and selective culture methods were employed. Samples were cultured on non-selective chocolate agar plates supplemented with IsoVitalX (bioMérieux SA) to identify the total aerobic bacteria. Chocolate agar is a rich culture medium used to identify the total aerobic bacteria (e.g., strictly aerobic, facultative aerobic-anaerobic and anaerobic-aerotolerant bacteria). Dilutions from 10^−1^ to 10^−7^ of each sample were prepared in 1X PBS, and 100 µL of the undiluted sample as well as 100 µL of each dilution were inoculated by spread plating. The plates were incubated for 24 h at 35 °C under aerobic conditions, then for 24 h at 22 °C under aerobic conditions.

The samples were simultaneously cultured on selective media. To isolate *Pseudomonas aeruginosa*, 20 μL of undiluted sample were spread plated on Cetrimide agar plates (bioMérieux SA). To isolate carbapenem-resistant *P. aeruginosa* and *Enterobacterales*, 20 μL of undiluted sample were spread plated on ChromID CARBA agar (bioMérieux SA) and ChromID OXA-48 agar plates (bioMérieux SA), respectively. To isolate glycopeptide-resistant *Enterococci*, 20 μL of undiluted sample were spread plated on a ChromID VRE agar plate (bioMérieux SA). Each plate was incubated for 48 hours at 35°C under aerobic conditions. All the colonies presenting different morphologies, as assessed visually from the selective and non-selective media, were enumerated and expressed as CFU/mL and identified using matrix-assisted laser desorption/ionization time-of-flight mass spectrometry (MALDI-TOF MS).

#### 2.3.2 Identification of isolates by MALDI-TOF MS

Spectral profiles were compared against the MBT IVD Library Revision J (2022, Bruker Daltonics), containing 4194 microbial reference spectra. Identification confidence was interpreted according to the manufacturer’s recommendations.

#### 2.3.3 Antimicrobial susceptibility testing

Phenotypic antimicrobial susceptibility testing was performed on the Enterobacterales and *P. aeruginosa* recovered from ChromID CARBA and ChromID OXA-48 selective media using the Kirby–Bauer disk diffusion method on Mueller–Hinton agar (MHA) and MHA supplemented with cloxacillin (MHA-cloxa) to facilitate AmpC β-lactamase detection (Jacoby, 2009), according to the French CA-SFM/EUCAST recommendations (“eucast: EUCAST,” n.d.). Bacterial suspensions were adjusted to a 0.5 McFarland standard (∼1.5 × 10⁸ CFU/mL) and uniformly spread onto agar plates. Standardized antibiotic discs were applied and plates incubated at 35°C for 18–24 h. Inhibition zone diameters were interpreted as susceptible (S), susceptible with increased exposure (I) or resistant (R) according to CA-SFM/EUCAST breakpoints.

#### 2.3.4 Detection of carbapenemase-producing strains

Phenotypic screening for CPE was performed according to European guidelines (“eucast: Clinical breakpoints and dosing of antibiotics,” n.d.) and as described previously (Hennebique et al., 2025). All s strains suspected of carbapenemase production were subsequently tested using the O.K.N.V.I. RESIST-5 immunochromatographic assay (Coris BioConcept), enabling detection of the five major families of carbapenemases: OXA-48, *Klebsiella pneumoniae* carbapenemase (KPC), New-Delhi metallo-β-lactamase (NDM), Verona integron–encoded metallo-β-lactamase (VIM) and imipenemase (IMP). Isolates negative by RESIST-5 but displaying compatible resistance phenotypes were further tested using the MAST CARBA PAcE assay (Mast Group). Confirmed carbapenemase-producing isolates were stored at −80°C in 20% glycerol until further analysis.

### 2.4 Clinical surveillance and patient isolates

#### 2.4.1 Surveillance screening and infection control measures

The infection prevention and control team at CHUGA adapted French national recommendations for the prevention of transmission of extensively drug-resistant (eXDR) bacteria for local use (Hennebique et al., 2025). Patients considered at risk of carrying eXDR bacteria were screened by rectal swab according to institutional procedures. Screening criteria included previous hospitalization abroad, known carriage of eXDR bacteria, transfer from another healthcare facility, residence in nursing homes, previous hospitalization, or documented contact with an eXDR carrier. Rectal swabs were inoculated on ChromID CARBA and ChromID OXA-48 agar plates (bioMérieux SA) for CPE sceening and on ChromID VRE plates (bioMérieux SA) for vancomycin-resistant *Enterococci* screening. Then, the colonies growing on these selective agar plates were screened in the same manner as for the environmental samples. Antibiotic susceptibility testing of the clinical strains was performed either by the disk diffusion method or by an automated system in liquid medium on a BD Phoenix™ M50. Hospitalized patients also undergo biological sampling for the diagnosis of bacterial infections (for example: blood culture, urine culture, respiratory specimen culture). When a bacterium isolated from a patient’s biological specimen was considered to be involved in the patient’s infection, an antibiotic susceptibility test was performed, and carbapenemase-producing strains were screened in the same ways as for rectal swabs.

#### 2.4.2 Clinical isolates and patient data

All adults hospitalized in the AICU and AIMCU between 1 January 2023 and 30 May 2024 were considered for inclusion. Patients with a CPE isolate identified during their ICU stay or previously known to carry a CPE were included in the study. According to French regulations, patient consent was not required for this retrospective study. The study was declared to the Data Protection Officer of CHUGA and reviewed by the Clinical Research Department of CHUGA. During the study period, 5,304 patients were hospitalized within the ICU building, including 1,241 patients in the medical AICU, 828 in the surgical AICU, 643 in the neurosurgical AICU, 994 in the medical AIMCU and 1,598 in the surgical AIMCU. Twenty-two CPE strains were identified during the study period. One CPE was excluded from genomic analyses because only a positive PCR result was available without strain isolation. Consequently, 21 clinical CPE isolates, coming from 18 patients, were included in comparative genomic analyses. These 21 strains included 18 strains isolated from rectal swabs, two strains isolated from blood culture and one strain isolated from a urine culture.

### 2.5 Whole-genome sequencing and bioinformatic analyses

#### 2.5.1 DNA extraction, sequencing and genome assembly

Genomic DNA was extracted from all environmental and clinical CPE isolates using the PureLink™ Genomic DNA Mini Kit (Invitrogen, Thermo Fisher Scientific), according to the manufacturer’s instructions for Gram-negative bacteria. DNA quantity and quality were assessed using a NanoDrop™ spectrophotometer (Thermo Fisher Scientific). Samples with a 260/280 ratio of 1.7–2.0 and DNA concentrations >20 ng/µL were retained for sequencing. Long-read WGS was performed by Plasmidsaurus (Houston, TX, USA) using Oxford Nanopore Technologies chemistry and R10.4.1 flow cells. Long-read sequencing data were assembled using the Plasmidsaurus assembly pipeline. Briefly, the lowest-quality 5% of reads were removed using Filtlong v0.2.1 (https://github.com/rrwick/Filtlong.git) with default parameters. Genome size was estimated using the helper functions implemented in Autocycler (Wick et al., 2025), and multiple read subsets were generated at optimal coverage levels. Each subset was assembled using three complementary long-read assemblers: Flye v2.9.6 (Kolmogorov et al., 2019), configured for high-quality Oxford Nanopore reads; Hifiasm (Cheng et al., 2021); and Plassembler v1.8.0 (Bouras et al., 2023) for plasmid detection and assembly. The resulting assemblies were processed using the Autocycler workflow, including the removal of low-depth and short contigs, compression, clustering, trimming, resolving and cleaning, followed by the generation of a consensus assembly. Consensus assemblies were rotated to an optimal starting position using dnaapler and polished with Medaka v1.8.0 (https://github.com/nanoporetech/medaka.git) using the filtered long reads. Short-read sequencing data were quality-filtered using fastp (Chen et al., 2018). Reads with Phred quality scores <15 or lengths <50 bp were discarded, and adapter and poly-G trimming were performed. Filtered short reads were aligned to the long-read assemblies using bwa-mem2, followed by polishing with Polypolish (Wick and Holt, 2022). For samples with sequencing coverage <25×, the --careful option was applied. Genome annotation was performed within the Plasmidsaurus pipeline using Bakta v1.11 and database v6.0 (Schwengers et al., 2021).

#### 2.5.2 Resistome, plasmid and taxonomic analyses

Acquired antimicrobial resistance genes were identified using ABRicate v1.2.0 (https://github.com/tseemann/abricate.git) with default parameters against the ResFinder database, accessed on 5 December 2025. Plasmids were reconstructed and classified using MOB-suite (Robertson and Nash, 2018) (mob_recon v3.1.9). MOB-suite classifications were used to distinguish plasmid-derived from chromosomal contigs and to support downstream plasmid analyses. Taxonomic assignments were independently assessed using GTDB-Tk v2.6.1 (Chaumeil et al., 2019) with the classify_wf workflow, based exclusively on chromosomal contigs. Genome completeness and contamination were evaluated using CheckM2 v1.0.1 (Chklovski et al., 2023). Average nucleotide identity (ANI) values were calculated on chromosomal contigs using FastANI v1.34 (Jain et al., 2018).

#### 2.5.3 Reassembly of selected isolates

To identify carbapenemase genes in isolates showing discrepancies between phenotypic and genomic carbapenemase detection, isolates 29, 30, 33, 34, 37, 39, 41, 50 and 57 were reassembled using Flye v2.9.6-b1802 (Kolmogorov et al., 2019) with the nano-hq option and polished using Medaka v2.2.1 (https://github.com/nanoporetech/medaka.git) with default parameters. The resulting assemblies were reanalysed using ResFinder and MOB-suite with the same parameters described above, and reconstructed plasmids were reannotated using Bakta v1.12 (Schwengers et al., 2021). Despite these analyses, the carbapenemase genes and associated plasmids could not be recovered for isolates 5, 8, 58 and 61, most likely because of low sequencing coverage or assembly limitations.

#### 2.5.4 Comparative genomic analyses

Comparative genomic analyses were performed using PIRATE v1.0.5 (Bayliss et al., 2019) with default parameters. GFF annotation files generated by the Plasmidsaurus pipeline using Bakta were used as input for pangenome analyses. Before analysis, contigs were separated into chromosomal and plasmid compartments according to MOB-suite classifications, and the two compartments were analysed independently. Newick trees generated from PIRATE outputs were used for plasmid phylogenetic analyses. Gene-context analyses of carbapenemase-carrying regions were performed using the annotated assemblies, and gene-context information extracted from the GFF files was visualised using the Interactive Tree Of Life (iTOL) platform (Letunic and Bork, 2021).

#### 2.5.5 Data availability

Raw sequencing reads and annotated assemblies are available from the European Nucleotide Archive under project PRJEB123823.

### 2.6 Statistical analyses

#### 2.6.1 Beta-diversity analysis of cultured bacterial communities

Beta diversity was assessed using Bray–Curtis dissimilarities calculated from the CFU/mL values of bacterial species cultured from WW and WWB samples. Differences in community composition between WW and WWB were tested by permutational multivariate analysis of variance (PERMANOVA) applied to the Bray– Curtis dissimilarity matrix, using 999 permutations. A P value <0.05 was considered indicative of a significant difference in beta diversity between the two groups. For visualisation, the distance of each sample to its respective group centroid was calculated and displayed using boxplots. No principal coordinate analysis (PCoA) was used for this visualisation.

#### 2.6.2 Association between carbapenemase genes and bacterial species

The distribution of carbapenemase genes (*bla_OXA-48_* and *bla_VIM_*) across bacterial species among environmental isolates (WW and WWB) was compared using the Fisher–Freeman–Halton exact test, an exact test of independence for contingency tables with more than two categories. Statistical significance was defined as P < 0.05.

#### 2.6.3 Temporal replacement of dominant environmental CPE populations

To test for temporal ecological replacement, the occurrence of Citrobacter spp. carrying blaOXA-48 was compared with that of Serratia spp. carrying blaVIM-4 across the sampling period (Supplementary Tables 1 and 5) using logistic regression. Isolates recovered from WW and WWB were combined for this analysis. Because of the small number of genomes available per time point, the significance of the slope term was assessed using a permutation test with 10,000 permutations. The two-sided permutation P value was calculated as the number of permuted slopes with an absolute value greater than or equal to the absolute observed slope, plus one, divided by the total number of permutations plus one (*n* + 1), where *n* = 10,000.

#### 2.6.4 Calculation of antimicrobial resistance scores

Antimicrobial susceptibility testing results were converted into numerical resistance scores, assigning susceptible (S), susceptible with increased exposure (I) and resistant (R) phenotypes values of 0, 0.5 and 1, respectively. For each isolate, a normalized resistance score was calculated as: Resistance score = Σ(R = 1, I = 0.5, S = 0) / N where N corresponds to the number of antibiotics tested for the isolate. Because environmental and clinical isolates were tested using different antimicrobial panels, resistance scores were calculated separately within each compartment. Pairwise comparisons were performed using Mann– Whitney U tests, whereas comparisons involving three carbapenemase groups were performed using Kruskal–Wallis tests. Statistical analyses and graphical representations were performed in Python v3.9.

#### 2.6.5 Comparison of additional antimicrobial resistance-gene counts

Acquired antimicrobial resistance genes were identified using ABRicate v1.2.0 against the ResFinder database, as described above. For each isolate with an annotated *blaOXA-48*, *blaVIM* or *blaNDM* gene, the number of additional acquired antimicrobial resistance genes was calculated after excluding the carbapenemase gene used for group assignment. Isolates for which no carbapenemase gene was recovered from the genomic assemblies were excluded from this analysis. Resistance-gene counts were compared pairwise between carbapenemase groups using two-sided Mann–Whitney U tests. A *P* value < 0.05 was considered statistically significant.

## Results

### 1. Diversity and temporal dynamics of cultured aerobic bacteria in AICU WW and WWBs

Over the 17-month study period (January 2023 to May 2024), 23 environmental samples (WW, *n* = 14; WWB, *n* = 9) were collected from the AICU WW outlet receiving effluent from the surgical and neurosurgical ICUs, the emergency department, and the mortuary (Hennebique et al., 2025). Culture-based microbiological analyses were performed to characterize cultivable aerobic bacterial communities and the occurrence of CPEs over time. WGS was performed for all confirmed CPE isolates, whereas non-CPE isolates were identified by MALDI-TOF MS only. CPE isolates initially identified as *Serratia marcescens* by MALDI-TOF MS were subsequently reassigned to *Serratia nevei* based on WGS and GTDB-Tk classification and are referred to as *S. nevei* throughout the manuscript.

Quantification of the cultivable aerobic bacterial community by serial dilution plating on chocolate agar yielded bacterial loads ranging from 4.3 × 10³ to 2.1 × 10⁶ CFU/mL. MALDI-TOF MS identified 25 cultivable bacterial taxa, predominantly Gram-negative species, whose composition varied over time and between WW and WWB (Fig. 1). Cultivable bacterial community composition differed significantly between WW and WWB, as determined by beta-diversity analysis based on Bray–Curtis dissimilarities (PERMANOVA, *P* <0.05; Supplementary Figure 1).

**Figure 1:**
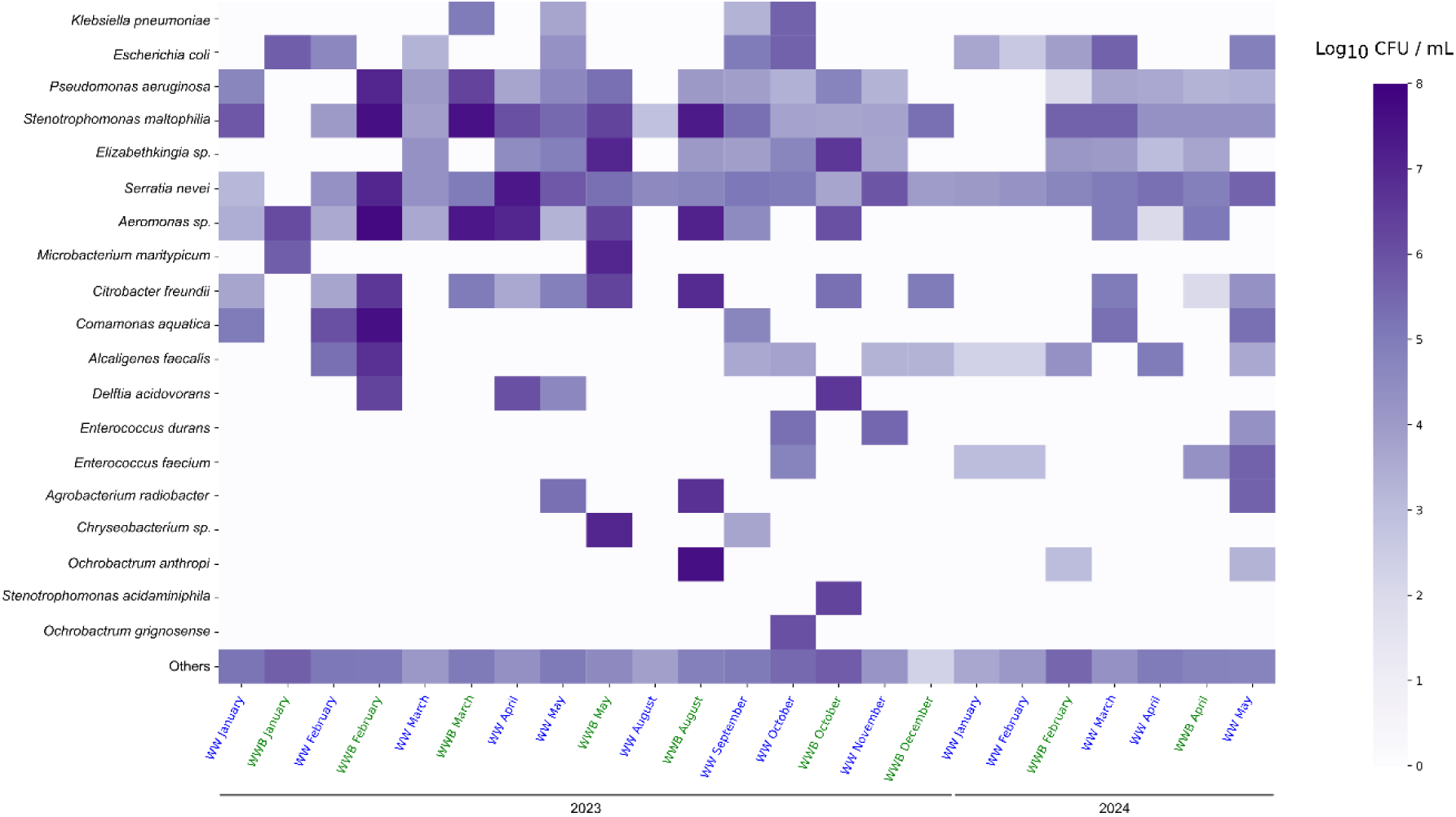
Temporal dynamics of cultivable bacterial abundance in WW and WWB from the AICU. Heatmap showing log₁₀-transformed colony-forming units per milliliter (CFU/mL) of total aerobic bacteria isolated from environmental samples collected over the study period (January 2023 to May 2024). Isolated bacterial strains were identified by MALDI-TOF-MS. CFUs/mL were determined as described in methods and expressed as log(10) per sink drain biofilm. Rows correspond to bacterial taxa identified by culture (species level when possible identified based on MALDI-TOF), and columns represent sampling time points. Color intensity reflects bacterial abundance, with darker shades indicating higher CFU/mL values.

Among the dominant taxa, the *C. freundii* complex and members of the *S. nevei* complex predominated. The *C. freundii* complex exhibited episodic increases in abundance, whereas members of the *S. nevei* complex were consistently recovered throughout the study period. Other opportunistic pathogens, including *Pseudomonas aeruginosa*, *Stenotrophomonas maltophilia*, and *Klebsiella pneumoniae*, were detected sporadically together with several low-abundance taxa.

### 2. Temporal shift in WW CPE population structure

After screening for the presence of CPEs, a total of 42 environmental CPE isolates were recovered from environmental (WW and WWB) samples across the entire study period (January 2023 to May 2024) with two principal carbapenemase families detected, namely *bla_OXA-48_* and *bla_VIM-4_*. Carbapenemase distribution differed significantly among bacterial species (Fisher–Freeman–Halton exact test, *P* < 0.001): *bla_OXA-48_* was predominantly associated with *Citrobacter spp*., whereas *bla_VIM-4_* was predominantly detected in *S. nevei*, and notably, no organisms contained both *bla_OXA-48_* and *bla_VIM-4_*. A marked ecological replacement was observed during the study period. While *bla_OXA-48_* -producing *Citrobacter* spp. predominated during the initial sampling phase, *bla_VIM-4_* -producing *S. nevei* progressively emerged and became the dominant environmental CPE population by the end of the study (Figure 2; logistic regression, effect of timepoint on *Citrobacter*(*bla_OXA-48_*) vs. *Serratia*(*bla_VIM-4_*) = −0.29, p = 0.0002). The structured distribution of resistance genes indicates that the observed population shift reflects both species turnover and a transition between distinct resistance gene reservoirs. The drivers underlying this shift remain unclear and were not specifically investigated in this study.

**Figure 2.**
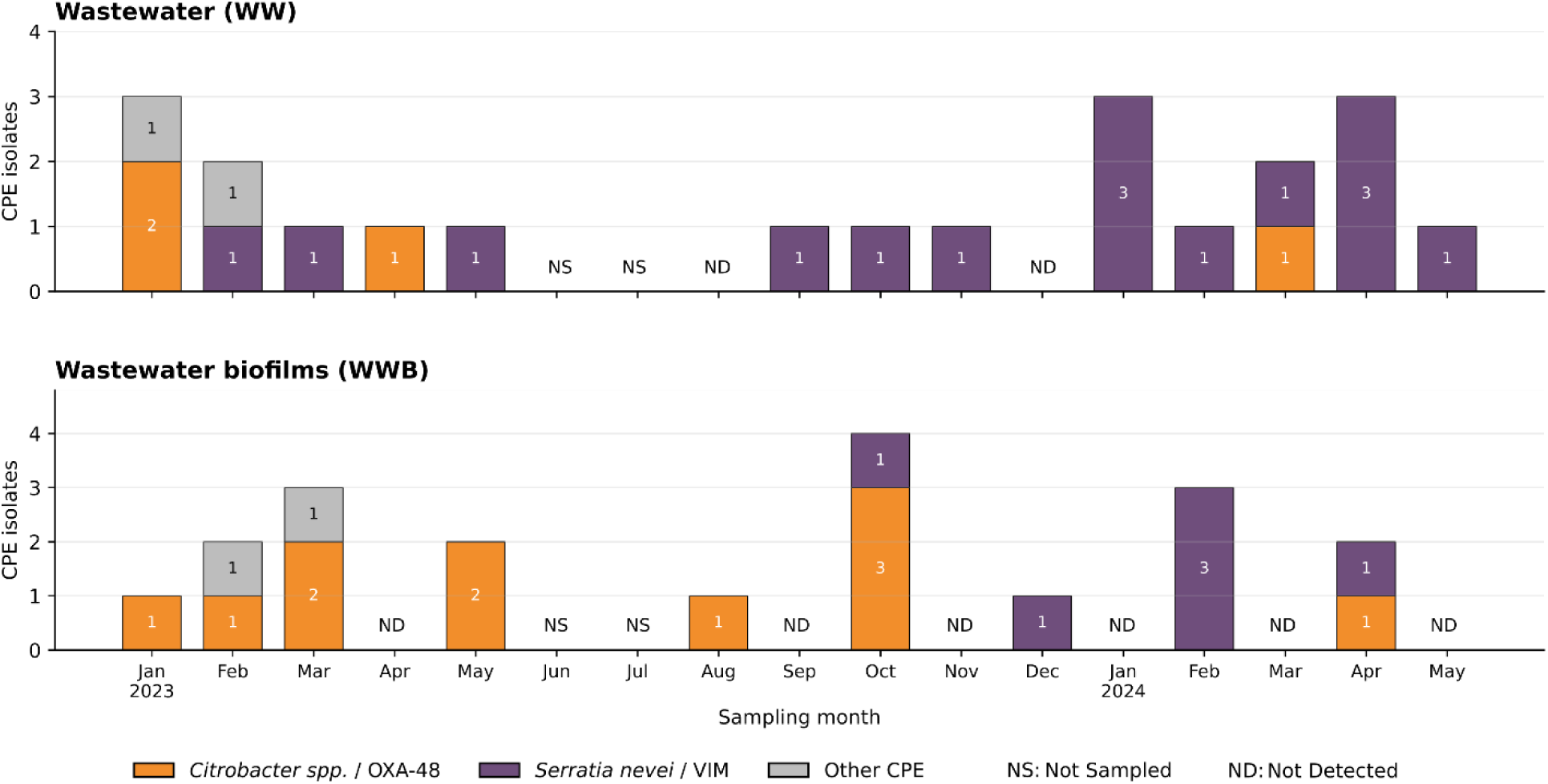
Temporal ecological dynamics of CPEs isolated from AICU WW and WWB. Temporal distribution of CPEs isolated from environmental samples between January 2023 and May 2024. The figure illustrates the progressive ecological replacement of *bla_OXA-48_* -producing *Citrobacter* spp. by *bla_VIM-4_* -producing *S. nevei* across WW and WWB compartments.

### 3. Phenotypic antimicrobial resistance profiles of environmental and clinical CPE isolates

Antimicrobial susceptibility profiles were compared among 42 environmental CPE isolates recovered from WW and WWB and 21 clinical CPE isolates recovered from patients hospitalized in the same ICU building between January 2023 and May 2024 (Supplementary Table 1). Antimicrobial susceptibility testing revealed extensive multidrug resistance among both environmental and clinical isolates (Supplementary Figure 2). Normalized resistance scores were calculated exclusively from phenotypic antimicrobial susceptibility testing results. Among environmental isolates, *bla_VIM-4_*-producing strains had significantly higher normalized resistance scores than *bla_OXA-48_* -producing strains in both WW (Mann–Whitney U test, P = 0.0053) and WWB (Mann–Whitney U test, P = 0.0003; Figure 3). Among clinical isolates, *bla_NDM_*-producing strains had the highest resistance scores, followed by *bla_VIM-4_*- and *bla_OXA-48_* -producing isolates. The overall difference among the three carbapenemase groups was significant (Kruskal–Wallis test, *P* = 0.0496). Dunn’s post hoc test with Holm correction identified a significant difference between *bla_VIM-4_*- and *bla_OXA-48_* - producing isolates (adjusted *P* = 0.0438), whereas the other pairwise comparisons were not significant.

**Figure 3.**
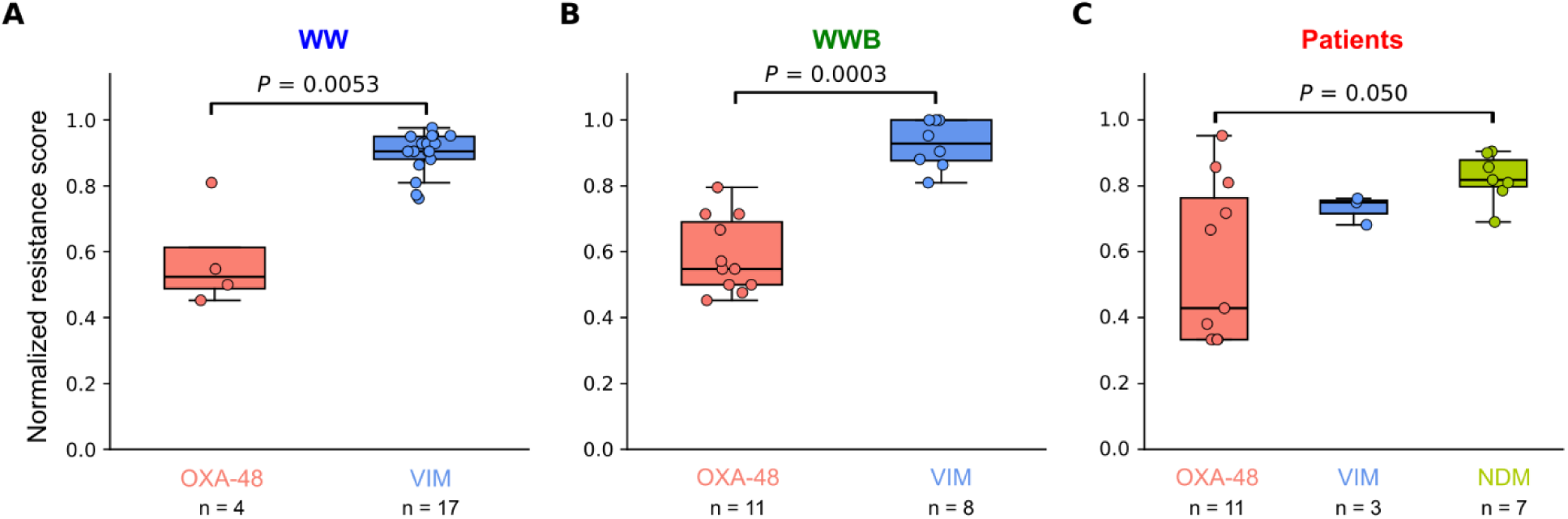
Phenotypic resistance profiles of CPEs from WW, WWB and hospitalized patients. (A) Normalized resistance scores of *bla_OXA-48_* - and *bla_VIM-4_*-producing isolates recovered from WW. (B) Normalized resistance scores of *bla_OXA-48_* - and *bla_VIM-4_*-producing isolates recovered from WWB. (C) Normalized resistance scores of *bla_OXA-48_* -, *bla_VIM-4_*- and *bla_NDM_*-producing clinical isolates. Resistance scores were calculated separately for each compartment to account for differences between the antimicrobial susceptibility testing panels used for environmental and clinical isolates. For each isolate, the normalized resistance score was calculated as the sum of the susceptibility scores across all antibiotics tested (R = 1, I = 0.5 and S = 0), divided by the total number of antibiotics tested (N). Boxes represent the interquartile range (IQR), horizontal lines indicate medians, whiskers extend to 1.5 × IQR, and dots represent individual isolates. *bla_OXA-48_* - and *bla_VIM-4_*-producing isolates were compared using two-sided Mann–Whitney U tests in WW (P = 0.0053) and WWB (P = 0.0003). Clinical carbapenemase groups were compared using a Kruskal–Wallis test (P = 0.0496). Exact P values are indicated in the figure.

Overall, environmental *bla_VIM-4_*- producing isolates displayed broader phenotypic resistance than *bla_OXA-48_* - producing isolates in both WW and WWB, whereas, *bla_NDM_*-producing clinical isolates exhibited the highest resistance scores among the clinical groups.

### 4. Genomic characterization of environmental and clinical CPE isolates

To disentangle chromosomal and plasmid contributions to carbapenemase dissemination, all 63 CPE isolates, comprising 42 environmental and 21 clinical isolates, were subjected to long-read WGS. Plasmid reconstruction and classification were performed using MOB-suite, allowing plasmid-derived and chromosomal contigs to be separated. Chromosomal sequences were used for species assignment, ANI calculations and core-genome clustering, whereas plasmid content, mobility and resistance-gene carriage were analysed independently. Genome quality metrics and ANI values are provided in Supplementary Tables 3 and 4.

Across all isolates, 94 plasmids carrying at least one antimicrobial resistance gene were identified, highlighting the contribution of plasmids to resistome organization. Resistance determinants were predominantly plasmid-borne, although chromosomal localization was also observed for several antimicrobial resistance gene classes and carbapenemase genes (Supplementary Figures 3 and 4). *bla*_OXA-48_ was mainly associated with IncL/M plasmids, whereas *bla*_VIM-4_ was predominantly associated with IncHI2A-rep1088 plasmids. In contrast, *bla*_NDM_ was identified on multiple plasmid backgrounds and on chromosomes. Beyond carbapenemases, plasmids frequently carried genes conferring resistance to aminoglycosides, sulfonamides, trimethoprim, quinolones and tetracyclines, resulting in complex multidrug-resistance platforms (Supplementary Figure 4 and Supplementary Table 5).

**Figure 4.**
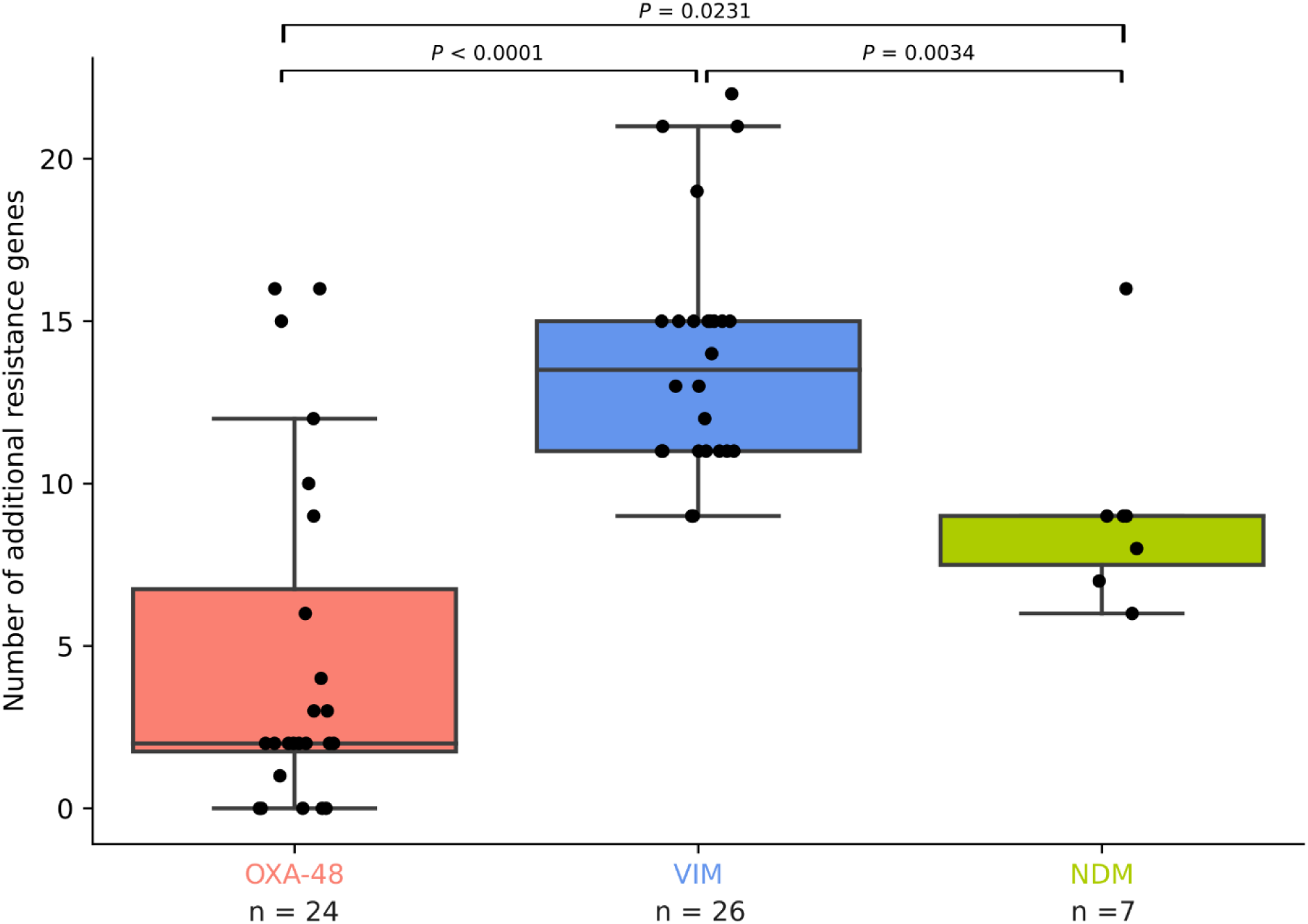
Additional ARGs in CPE isolates carrying *bla_OXA-48_*, *bla_VIM_* or *bla_NDM_*. Boxplots show the number of antimicrobial resistance genes identified in each isolate, excluding the carbapenemase gene used for group assignment. Each dot represents an individual isolate, and the number of isolates in each group is indicated below the x-axis. Boxes represent the interquartile range, horizontal lines indicate medians, and whiskers extend to 1.5 times the interquartile range. Pairwise comparisons between carbapenemase groups were performed using two-sided Mann–Whitney U tests. *P* values are indicated above the corresponding comparisons.

Phenotypic carbapenemase profiles showed high overall concordance with genomic predictions. Initial discrepancies between phenotypic and genomic carbapenemase detection were observed for 11 isolates and were largely resolved following reassembly and plasmid reconstruction. However, for isolates 5, 8, 58 and 61, the carbapenemase genes and associated plasmids could not be recovered from the genomic assemblies, likely because of low sequencing coverage or assembly limitations.

The number of additional acquired antimicrobial resistance genes was compared among the 57 isolates with an annotated OXA-48, VIM or NDM carbapenemase gene (Figure 4). VIM-carrying isolates contained significantly more additional resistance genes than OXA-48-carrying isolates (two-sided Mann–Whitney U test, *P* <0.0001) and NDM-carrying isolates (*P* = 0.0034). NDM-carrying isolates also contained significantly more additional resistance genes than OXA-48-carrying isolates (*P* = 0.0231). These genomic results were consistent with the broader phenotypic resistance observed among VIM-producing isolates and the comparatively high resistance scores of NDM-producing clinical isolates.

### 5. Dominant carbapenemase dissemination platforms across environmental and clinical compartments

Carbapenemase distribution was strongly structured according to both bacterial host and compartment. *bla_OXA-48_* -producing isolates were predominantly associated with *Citrobacter* spp., particularly within WW and WWB compartments, whereas *bla_OXA-48_* -producing *E. coli* isolates were mainly recovered from patients. In contrast, *bla_VIM-4_* -producing isolates were dominated by *S. nevei*, which accounted for 21 of 28 *bla_VIM-4_* - positive isolates and was exclusively detected in environmental compartments. *bla*_NDM_-producing isolates were recovered exclusively from patients and were primarily associated with *E. intestinihominis* (Figure 5a). Among the 26 *bla_OXA-48_* -producing isolates, 21 (81%) carried plasmid-borne carbapenemase genes, whereas five (19%) harboured chromosomal copies. Pangenome based plasmid clustering using PIRATE together with comparative analysis of the *bla_OXA-48_* genetic contexts revealed that the majority of plasmid associated *bla_OXA-48_* genes were located on highly conserved IncL/M plasmid backbones, indicating that these plasmid backbones are shared across environmental and clinical compartments and diverse host species (Figure 5B and 5C; Supplementary Figure 5). Similarly, 25 of 28 (89%) *bla_VIM-4_* -producing isolates carried plasmid-borne *bla_VIM-4_* −4 genes, while only one isolate (4%) harboured a chromosomal copy, while for two isolates (sample number five and eight) the *bla_VIM_* could not be recovered by ONT sequencing. The majority of *bla_VIM-4_*-associated plasmids belonged to the IncHI2A-rep1088 group, whereas only a small number were associated with alternative plasmid backgrounds (Figure 5B; Supplementary Figure 5). In addition, *bla_VIM-4_* was consistently embedded within a highly conserved class 1 integron-associated genetic structure, further supporting the persistence of a dominant IncHI2A-associated dissemination platform (Figure 5C). Although IncHI2A-rep1088 plasmids carrying *bla_VIM-4_* were predominantly recovered from *S. nevei*, the same MOB-suite plasmid cluster was identified in isolate 13, a *C. freundii* isolate recovered from WWB in March 2023. This cross-species occurrence, together with the similarity of the plasmid backbones and carbapenemase-gene contexts, is consistent with dissemination of this plasmid background between bacterial hosts. In contrast, *bla*_NDM_-producing isolates displayed heterogeneous genetic backgrounds involving multiple plasmid types and chromosomal locations. Chromosomal integration was observed in five of seven *bla*_NDM_-producing isolates (71%), and no dominant dissemination platform was identified (Figure 5C; Supplementary Figure 5).

**Figure 5.**
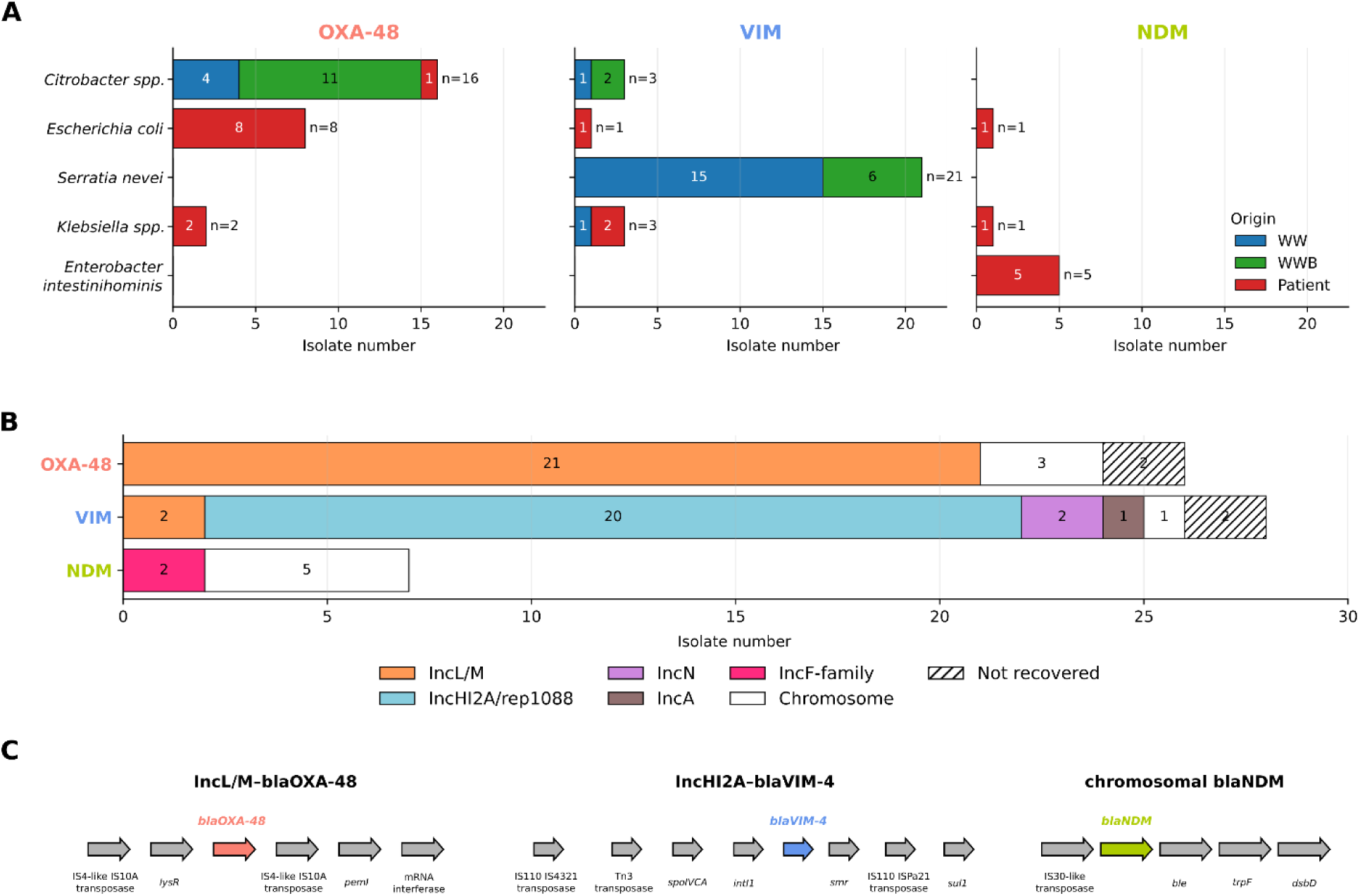
Shared carbapenemase plasmid backbones across environmental and clinical compartments. **(A)** Distribution of *bla_OXA-48_* -, *bla_VIM-4_* - and NDM-producing isolates according to bacterial host and compartment. WW, WWB and patient isolates are shown in blue, green and red, respectively. **(B)** Genetic support of carbapenemase genes. Bars indicate the number of isolates carrying carbapenemase genes on different plasmid backgrounds or chromosomes. *bla_OXA-48_* was predominantly associated with IncL/M plasmids, whereas *bla_VIM-4_* was mainly associated with IncHI2A-rep1088 plasmids. In contrast, NDM occurred on multiple plasmid backgrounds and chromosomal locations. **(C)** Representative genetic environments surrounding *bla_OXA-48_*, *bla_VIM-4_* and *bla_NDM_* identified in the isolate collection. *bla_OXA-48_* and *bla_VIM-4_* were associated with highly conserved genetic structures, whereas NDM occurred in more diverse genomic contexts. Representative structures correspond to the most frequently observed arrangements within each carbapenemase group. Numbers within bars indicate isolate counts.

## Discussion

We combined longitudinal CPE surveillance of WW and WWB with comparative WGS of environmental isolates and contemporaneous clinical isolates recovered from patients hospitalized in the same hospital building connected to the studied WW outlet. This approach enabled us to investigate the ecological dynamics of CPE bacterial hosts together with the genomic organization of their carbapenemase carrying genetic elements across environmental and clinical compartments.

Our longitudinal analysis revealed substantial temporal changes in the dominant environmental bacterial hosts carrying carbapenemases, with *bla_OXA-48_*-producing *Citrobacter* spp. progressively replaced by *bla_VIM-4_*-producing *S. nevei* over the course of the study. This succession illustrates that dominant environmental CPE populations can change substantially over relatively short timescales. The expansion of *S. nevei* may have been influenced by species-specific ecological traits. Type VI secretion systems, which are widespread among *Serratia* spp., mediate antagonistic interactions with neighbouring bacteria and could potentially provide a competitive advantage in densely populated biofilm communities (Jiang et al., 2025). However, this mechanism was not investigated in the present study and remains to be experimentally tested. Although previous studies have documented CPE persistence in hospital WW and associated biofilms (Koh et al., 2025; Mollenkopf et al., 2025; Schussman et al., n.d.) ([reference 1](https://pmc.ncbi.nlm.nih.gov/articles/PMC12889080/); [reference 2](https://www.sciencedirect.com/science/article/pii/S0048969725008769); [reference 3](https://pmc.ncbi.nlm.nih.gov/articles/PMC12569050/#Sec10)), temporal changes in the dominant bacterial hosts carrying different carbapenemases have received comparatively limited attention. Our findings therefore emphasize the importance of longitudinal surveillance for detecting ecological succession and shifts in dominant host–carbapenemase associations that would remain undetected in cross-sectional studies.

The contrasting organization of environmental and clinical CPE populations was particularly striking. Whereas the environmental population was successively dominated by two host–carbapenemase associations—*Citrobacter* spp.– *bla_OXA-48_* and *S. nevei*– *bla_VIM-4_*—clinical isolates remained taxonomically diverse throughout the study period. This contrast suggests that conditions within WW and WWB favour the persistence and expansion of specific bacterial hosts, whereas the clinical reservoir reflects the repeated introduction of diverse CPE lineages. Biofilm formation is likely to contribute to this ecological structuring by providing protected niches that promote long-term bacterial persistence and facilitate HGT, consistent with previous studies identifying hospital drain biofilms as persistent reservoirs of CPE (Hennebique et al., 2025; Park et al., 2020; Stoesser et al., 2024). Consistent with this hypothesis, WWB exhibited greater species diversity and supported the persistence of several low abundance taxa compared with planktonic WW (Figure 1 and 2).

Comparative plasmid genomics revealed a central feature of carbapenemase dissemination in this system: the observed patterns were structured around a limited number of closely related plasmid platforms recovered from phylogenetically distinct Enterobacterales species, rather than being explained solely by the expansion of individual bacterial lineages. This was particularly evident for *bla_OXA-48_*, which was predominantly carried by closely related IncL/M plasmid backbones recovered from different bacterial hosts and across environmental and clinical compartments. Similarly, *bla_VIM-4_*- was mainly embedded within a shared class 1 integron-associated genetic context on IncHI2A-rep1088 plasmids recovered from environmental isolates (Figure 5 and Supplementary Figure 5). The repeated recovery of these closely related carbapenemase-associated platforms across sampling times, bacterial hosts and compartments indicates that successful plasmid backbones can persist across species boundaries and contribute to carbapenemase dissemination independently of the dominant bacterial host. Thus, bacterial populations and carbapenemase-carrying genetic elements followed partly distinct ecological trajectories within the hospital WW ecosystem.

In contrast, *bla_NDM_* -producing isolates displayed a more heterogeneous genomic organization and was detected on different genetic supports, including IncF-family plasmids and chromosomal locations (Lerminiaux et al., 2025; Sakamoto et al., 2022). This pattern is noteworthy because *bla_NDM_* is generally described as plasmid-associated in Enterobacterales, with IncX3, IncF, IncC, IncN and IncHI-type plasmids frequently implicated in its dissemination, whereas chromosomal *bla_NDM_* integration has been reported less commonly (Luo et al., 2021). In our collection, chromosomal *bla_NDM_* was mainly observed in patient-derived *Enterobacter intestinihominis* isolates, likely reflecting the expansion of a related clinical lineage rather than repeated acquisition of diverse *bla_NDM_* plasmids.

Beyond carbapenemases, the detected plasmids also frequently carried genes conferring resistance to aminoglycosides, sulfonamides, trimethoprim, tetracyclines and quinolones, demonstrating that they represent multidrug-resistance vehicles rather than simple carbapenemase carriers. *bla_VIM-4_*-carrying isolates contained significantly more additional ARGs than *bla_OXA-48_*- or *bla_NDM_* -carrying isolates, while *bla_NDM_* -carrying isolates contained significantly more ARGs than *bla_OXA-48_*-carrying isolates (Figure 4). These genomic differences paralleled the phenotypic resistance profiles: environmental *bla_VIM_* -producing isolates exhibited significantly higher resistance scores than *bla_OXA-48_*-producing isolates, while *bla_NDM_* -producing isolates displayed the highest resistance scores among the clinical groups (Figure 3). Together, these findings demonstrate an association between a greater acquired resistance-gene burden and broader phenotypic resistance across carbapenemase groups. Co-selection by non-carbapenem antibiotics, disinfectants and other environmental stressors may therefore promote the persistence of these multidrug-resistance platforms, even in the absence of direct carbapenem exposure (Bengtsson-Palme et al., 2018; Gillings, 2013; Lee et al., 2017), and may contribute to their repeated recovery within hospital WW communities.

A major strength of this study lies in the analytical framework used to disentangle bacterial and plasmid evolution. By combining comparative WGS, MOB-suite-based plasmid reconstruction, independent chromosomal taxonomic assignment, ANI and pangenome analyses, bacterial population dynamics and carbapenemase carrying genetic elements could be investigated separately. This approach minimized biases associated with plasmid-derived sequences during species assignment and enabled direct comparison of chromosomal and plasmid evolutionary patterns. More broadly, integrating longitudinal environmental surveillance with comparative genomics provides a robust framework for investigating antimicrobial resistance ecology across interconnected environmental and clinical reservoirs.

Several limitations should nevertheless be acknowledged. First, this study was conducted within a single hospital WW network, and the ecological and genomic dynamics described here may differ among healthcare facilities depending on patient populations, antimicrobial usage and WW infrastructure. Second, only cultivable CPEs were investigated, potentially overlooking uncultured or low abundance populations. Third, although complete plasmid reconstruction enabled detailed characterization of carbapenemase carrying genetic elements, our study cannot distinguish horizontal plasmid transfer from clonal expansion as the primary mechanism underlying the observed distribution of conserved plasmid backbones. Future studies integrating metagenomics, metatranscriptomics and experimental approaches to quantify plasmid transfer within wastewater biofilms will help resolve these processes and further improve our understanding of antimicrobial resistance persistence in hospital WW.

Overall, our findings demonstrate that the bacterial hosts carrying carbapenemase genes can change markedly over time, whereas the genetic vehicles carrying those genes remain comparatively conserved. Distinguishing the ecological dynamics of bacterial hosts from those of carbapenemase carrying genetic elements provides a more comprehensive understanding of antimicrobial resistance persistence in hospital WW. More broadly, our findings highlight the importance of integrating longitudinal environmental surveillance with comparative genomics to understand the ecology of clinically important carbapenemase genes and support the incorporation of environmental genomic surveillance into One Health strategies for antimicrobial resistance monitoring.

## Conclusions

- Longitudinal surveillance revealed temporal changes in the dominant environmental CPE bacterial hosts.
- Despite the ecological turnover of dominant bacterial hosts, comparative genomics identified highly similar IncL/M plasmid backbones carrying *bla_OXA-48_* and related *bla_VIM-4_* -associated genetic contexts. Clinical CPE populations remained taxonomically diverse and did not mirror environmental succession.
- Distinguishing bacterial host dynamics from carbapenemase-carrying genetic elements provides a more comprehensive understanding of CPE persistence in hospital wastewater ecosystems.
- Integrating longitudinal surveillance with comparative genomics can strengthen environmental surveillance of clinically relevant antimicrobial resistance.

## Author contributions (CRediT)

**Morgane Roger-Margueritat:** Investigation, Methodology, Formal analysis, Data curation, Visualization, Writing – review & editing.

**Verena Schmidt:** Investigation, Formal analysis, Data curation, Validation, Writing – review & editing.

**Gregory E. McCallum:** Software, Formal analysis, Visualization, Writing – review & editing.

**Eva Gendron:** Investigation, Data curation, Formal analysis.

**Jostin Monge-Ruiz:** Investigation, Data curation.

**Patrice Morand:** Resources.

**Claire Terreaux-Masson:** Investigation.

**Caroline Landelle:** Conceptualization, Methodology, Resources, Supervision, Writing – review & editing.

**James P. J. Hall:** Formal analysis, Writing – review & editing.

**Aurélie Hennebique:** Conceptualization, Formal analysis, Methodology, Validation, Resources, Supervision, Writing – review & editing.

**Elena Buelow:** Conceptualization, Methodology, Investigation, Formal analysis, Validation, Project administration, Supervision, Funding acquisition, Writing – original draft, Writing – review & editing.

All authors read and approved the final manuscript.

## References

Abe, K., Nomura, N., Suzuki, S., 2020. Biofilms: hot spots of horizontal gene transfer (HGT) in aquatic environments, with a focus on a new HGT mechanism. FEMS Microbiol. Ecol. 96. https://doi.org/infecti

Anantharajah, A., Goormaghtigh, F., Nguvuyla Mantu, E., Güler, B., Bearzatto, B., Momal, A., Werion, A., Hantson, P., Kabamba-Mukadi, B., Van Bambeke, F., Rodriguez-Villalobos, H., Verroken, A., 2024. Long-term intensive care unit outbreak of carbapenemase-producing organisms associated with contaminated sink drains. J. Hosp. Infect. 143, 38–47. 10.1016/j.jhin.2023.10.010

Arredondo-Alonso, S., Willems, R.J., van Schaik, W., Schürch, A.C., 2017. On the (im)possibility of reconstructing plasmids from whole-genome short-read sequencing data. Microb. Genomics 3, e000128. 10.1099/mgen.0.000128

Bayliss, S.C., Thorpe, H.A., Coyle, N.M., Sheppard, S.K., Feil, E.J., 2019. PIRATE: A fast and scalable pangenomics toolbox for clustering diverged orthologues in bacteria. GigaScience 8, giz119. 10.1093/gigascience/giz119

Bengtsson-Palme, J., Kristiansson, E., Larsson, D.G.J., 2018. Environmental factors influencing the development and spread of antibiotic resistance. FEMS Microbiol. Rev. 42, 10.1093/femsre/fux053. https://doi.org/10.1093/femsre/fux053

Bortolaia, V., Kaas, R.S., Ruppe, E., Roberts, M.C., Schwarz, S., Cattoir, V., Philippon, A., Allesoe, R.L., Rebelo, A.R., Florensa, A.F., Fagelhauer, L., Chakraborty, T., Neumann, B., Werner, G., Bender, J.K., Stingl, K., Nguyen, M., Coppens, J., Xavier, B.B., Malhotra-Kumar, S., Westh, H., Pinholt, M., Anjum, M.F., Duggett, N.A., Kempf, I., Nykäsenoja, S., Olkkola, S., Wieczorek, K., Amaro, A., Clemente, L., Mossong, J., Losch, S., Ragimbeau, C., Lund, O., Aarestrup, F.M., 2020. ResFinder 4.0 for predictions of phenotypes from genotypes. J. Antimicrob. Chemother. 75, 3491–3500. 10.1093/jac/dkaa345

Bouras, G., Sheppard, A.E., Mallawaarachchi, V., Vreugde, S., 2023. Plassembler: an automated bacterial plasmid assembly tool. Bioinformatics 39, btad409. 10.1093/bioinformatics/btad409

Buelow, E., Dauga, C., Carrion, C., Mathé-Hubert, H., Achaibou, S., Gaschet, M., Jové, T., Chesneau, O., Kennedy, S.P., Ploy, M.-C., Da Re, S., Dagot, C., 2023. Hospital and urban wastewaters shape the matrix and active resistome of environmental biofilms. Water Res. 244, 120408. 10.1016/j.watres.2023.120408

Chaumeil, P.-A., Mussig, A.J., Hugenholtz, P., Parks, D.H., 2019. GTDB-Tk: a toolkit to classify genomes with the Genome Taxonomy Database. Bioinformatics 36, 1925–1927. 10.1093/bioinformatics/btz848

Chen, S., Zhou, Y., Chen, Y., Gu, J., 2018. fastp: an ultra-fast all-in-one FASTQ preprocessor. Bioinformatics 34, i884–i890. 10.1093/bioinformatics/bty560

Cheng, H., Concepcion, G.T., Feng, X., Zhang, H., Li, H., 2021. Haplotype-resolved de novo assembly using phased assembly graphs with hifiasm. Nat. Methods 18, 170–175. 10.1038/s41592-020-01056-5

Chklovski, A., Parks, D.H., Woodcroft, B.J., Tyson, G.W., 2023. CheckM2: a rapid, scalable and accurate tool for assessing microbial genome quality using machine learning. Nat. Methods 20, 1203– 1212. 10.1038/s41592-023-01940-w

De Geyter, D., Blommaert, L., Verbraeken, N., Sevenois, M., Huyghens, L., Martini, H., Covens, L., Piérard, D., Wybo, I., 2017. The sink as a potential source of transmission of carbapenemase-producing Enterobacteriaceae in the intensive care unit. Antimicrob. Resist. Infect. Control 6, 24. 10.1186/s13756-017-0182-3

eucast: Clinical breakpoints and dosing of antibiotics [WWW Document], n.d. URL https://www.eucast.org/clinical_breakpoints (accessed 4.7.25).

eucast: EUCAST [WWW Document], n.d. URL https://www.eucast.org/ (accessed 10.17.24).

Gillings, M.R., 2013. Evolutionary consequences of antibiotic use for the resistome, mobilome and microbial pangenome. Front. Microbiol. 4, 4. 10.3389/fmicb.2013.00004

Hennebique, A., Monge-Ruiz, J., Roger-Margueritat, M., Morand, P., Terreaux-Masson, C., Maurin, M., Mercier, C., Landelle, C., Buelow, E., 2025. The hospital sink drain biofilm resistome is independent of the corresponding microbiota, the environment and disinfection measures. Water Res. 284, 123902. 10.1016/j.watres.2025.123902

Jacoby, G.A., 2009. AmpC beta-lactamases. Clin. Microbiol. Rev. 22, 161–182, Table of Contents. 10.1128/CMR.00036-08

Jain, C., Rodriguez-R, L.M., Phillippy, A.M., Konstantinidis, K.T., Aluru, S., 2018. High throughput ANI analysis of 90K prokaryotic genomes reveals clear species boundaries. Nat. Commun. 9, 5114. 10.1038/s41467-018-07641-9

Jiang, L., Yi, W., Zhao, Y., Zhu, N., Zhao, D., Peng, Z., Song, L., Dong, T., Jiang, X., Liu, D., Ji, X., Guan, Q., Jiang, H., 2025. Comprehensive genomic analysis of type VI secretion system diversity and associated proteins in *Serratia*. Microb. Genomics 11, 001424. 10.1099/mgen.0.001424

Koh, V., Cabrera, R., Sridatta, P.S.R., Thevasagayam, N.M., Lim, Z.Q., Marimuthu, K., Venkatachalam, I., Cherng, B.P.Z., Fong, R.K.C., Pada, S.K., Ooi, S.T., Smitasin, N., Thoon, K.C., Hsu, L.Y., Koh, T.H., De, P.P., Tan, T.Y., Chan, D., Deepak, R.N., Tee, N.W.S., Gan, Y.-H., Matlock, W., Eyre, D.W., Ang, M., Lin, R.T.P., Teo, J., Ng, O.T., 2025. Plasmid dynamics driving carbapenemase gene dissemination in healthcare environments: a nationwide analysis of closed Enterobacterales genomes. Nat. Commun. 16, 9522. 10.1038/s41467-025-64515-7

Kolmogorov, M., Yuan, J., Lin, Y., Pevzner, P.A., 2019. Assembly of long, error-prone reads using repeat graphs. Nat. Biotechnol. 37, 540–546. 10.1038/s41587-019-0072-8

Lee, J., Jeon, J.H., Shin, J., Jang, H.M., Kim, S., Song, M.S., Kim, Y.M., 2017. Quantitative and qualitative changes in antibiotic resistance genes after passing through treatment processes in municipal wastewater treatment plants. Sci. Total Environ. 605-606, 906–914. 10.1016/j.scitotenv.2017.06.250

Lerminiaux, N., Fakharuddin, K., Longtin, Y., McGill, E., Mitchell, R., Mataseje, L., On Behalf Of The Canadian Nosocomial Infection Surveillance Program, null, 2025. Plasmid genomic epidemiology of bla NDM carbapenemase-producing Enterobacterales in Canada from 2010 to 2023. Microb. Genomics 11, 001415. 10.1099/mgen.0.001415

Letunic, I., Bork, P., 2021. Interactive Tree Of Life (iTOL) v5: an online tool for phylogenetic tree display and annotation. Nucleic Acids Res. 49, W293–W296. 10.1093/nar/gkab301

Logan, L.K., Weinstein, R.A., 2017. The Epidemiology of Carbapenem-Resistant Enterobacteriaceae: The Impact and Evolution of a Global Menace. J. Infect. Dis. 215, S28–S36. 10.1093/infdis/jiw282

Ludden, C., Reuter, S., Judge, K., Gouliouris, T., Blane, B., Coll, F., Naydenova, P., Hunt, M., Tracey, A., Hopkins, K.L., Brown, N.M., Woodford, N., Parkhill, J., Peacock, S.J., 2017. Sharing of carbapenemase-encoding plasmids between Enterobacteriaceae in UK sewage uncovered by MinION sequencing. Microb. Genomics 3, e000114. 10.1099/mgen.0.000114

Luo, X., Yin, Z., Zeng, L., Hu, L., Jiang, X., Jing, Y., Chen, F., Wang, D., Song, Y., Yang, H., Zhou, D., 2021. Chromosomal Integration of Huge and Complex *blaNDM*-Carrying Genetic Elements in Enterobacteriaceae. Front. Cell. Infect. Microbiol. 11. 10.3389/fcimb.2021.690799

M, M., C, S., N, M.S., A, K., L, O., N, C., B, H., N, D., W, B., G, D., M, C., D, M., Sc, C., G, M., B, G.-Z., Lp, B., 2025. Spatiotemporal and genomic analysis of carbapenem resistance elements in Enterobacterales from hospital inpatients and natural water ecosystems of an Irish city. Microbiol. Spectr. 13. 10.1128/spectrum.00904-24

Mathers, A.J., Cox, H.L., Kitchel, B., Bonatti, H., Brassinga, A.K.C., Carroll, J., Scheld, W.M., Hazen, K.C., Sifri, C.D., 2011. Molecular dissection of an outbreak of carbapenem-resistant enterobacteriaceae reveals Intergenus KPC carbapenemase transmission through a promiscuous plasmid. mBio 2, e00204–00211. 10.1128/mBio.00204-11

Mathers, A.J., Li, T.J.X., He, Q., Narendra, S., Stoesser, N., Eyre, D.W., Walker, A.S., Barry, K.E., Castañeda-Barba, S., Huang, F.W., Parikh, H., Kotay, S., Crook, D.W., Reidys, C., 2024. Developing a framework for tracking antimicrobial resistance gene movement in a persistent environmental reservoir. Npj Antimicrob. Resist. 2, 50. 10.1038/s44259-024-00069-w

Mollenkopf, D.F., Lee, S., Ballash, G.A., Sulliván, S.M.P., Lee, J., Wittum, T.E., 2025. Carbapenemase-producing Enterobacterales and their carbapenemase genes are stably recovered across the wastewater-watershed ecosystem nexus. Sci. Total Environ. 975, 179241. 10.1016/j.scitotenv.2025.179241

Park, S.C., Parikh, H., Vegesana, K., Stoesser, N., Barry, K.E., Kotay, S.M., Dudley, S., Peto, T.E.A., Crook, D.W., Walker, A.S., Mathers, A.J., 2020. Risk Factors Associated with Carbapenemase-Producing Enterobacterales (CPE) Positivity in the Hospital Wastewater Environment. Appl. Environ. Microbiol. 86, e01715–20. 10.1128/AEM.01715-20

Partridge, S.R., Kwong, S.M., Firth, N., Jensen, S.O., 2018. Mobile Genetic Elements Associated with Antimicrobial Resistance. Clin. Microbiol. Rev. 31, e00088–17. 10.1128/CMR.00088-17

Robertson, J., Nash, J.H.E., 2018. MOB-suite: software tools for clustering, reconstruction and typing of plasmids from draft assemblies. Microb. Genomics 4, e000206. 10.1099/mgen.0.000206

Sakamoto, N., Akeda, Y., Sugawara, Y., Matsumoto, Y., Motooka, D., Iida, T., Hamada, S., 2022. Role of Chromosome- and/or Plasmid-Located blaNDM on the Carbapenem Resistance and the Gene Stability in Escherichia coli. Microbiol. Spectr. 10, e0058722. 10.1128/spectrum.00587-22

Schussman, M.K., Feng, S., Schmoldt, A., Bootsma, M.J., Dennis, K., Janssen, K.H., McLellan, S.L., n.d. Environmental reservoirs account for high levels of carbapenem resistance genes in wastewater. Microbiol. Spectr. 14, e01737–25. 10.1128/spectrum.01737-25

Schwengers, O., Jelonek, L., Dieckmann, M.A., Beyvers, S., Blom, J., Goesmann, A., 2021. Bakta: rapid and standardized annotation of bacterial genomes via alignment-free sequence identification. Microb. Genomics 7, 000685. 10.1099/mgen.0.000685

Stoesser, N., George, R., Aiken, Z., Phan, H.T.T., Lipworth, S., Quan, T.P., Mathers, A.J., De Maio, N., Seale, A.C., Eyre, D.W., Vaughan, A., Swann, J., Peto, T.E.A., Crook, D.W., Cawthorne, J., Dodgson, A., Walker, A.S., TRACE Investigators Group, 2024. Genomic epidemiology and longitudinal sampling of ward wastewater environments and patients reveals complexity of the transmission dynamics of bla KPC-carbapenemase-producing Enterobacterales in a hospital setting. JAC-Antimicrob. Resist. 6, dlae140. 10.1093/jacamr/dlae140

Wick, R.R., Holt, K.E., 2022. Polypolish: Short-read polishing of long-read bacterial genome assemblies. PLoS Comput. Biol. 18, e1009802. 10.1371/journal.pcbi.1009802

Wick, R.R., Howden, B.P., Stinear, T.P., 2025. Autocycler: long-read consensus assembly for bacterial genomes. Bioinformatics 41, btaf474. 10.1093/bioinformatics/btaf474

