## Supplementary Figures for "Distinct bacterial hosts, shared *bla_OXA-48_* plasmid backbones: longitudinal comparative genomics of carbapenemase-producing Enterobacterales from hospital wastewater, biofilms and patients"

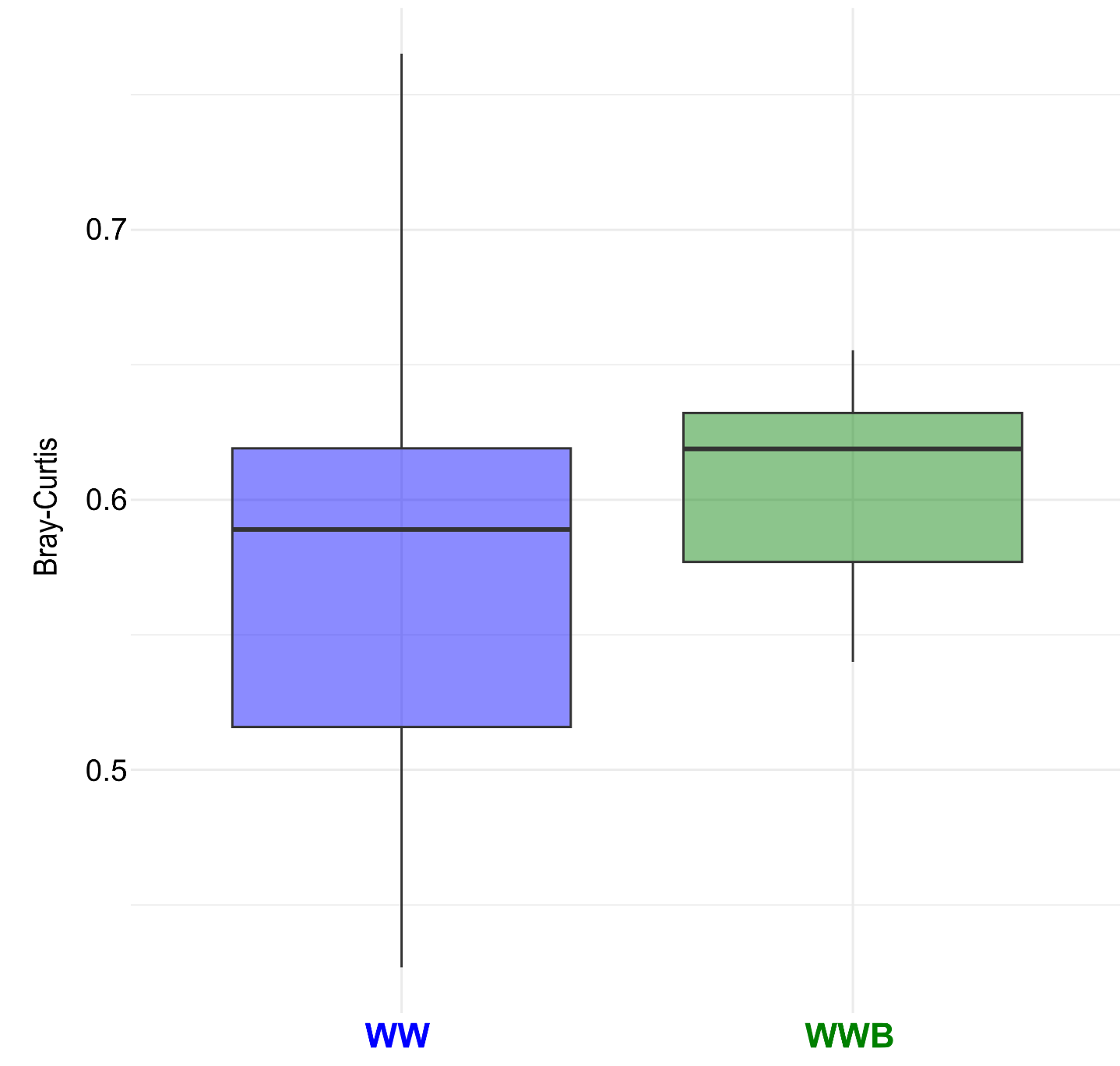

**Supplementary Figure 1**: Boxplot visualizing the beta-diversity via Bray-Curtis distances of the microbiota of WW (blue) and WWB (green) determined by CFU/mL. Depicted taxa are on species level. The distance to centroid for each sample is used to make the boxplot. Permanova was used with 999 permutations on distances between WW and WWB and a P value < 0.05 indicates a significant beta-diversity difference between both groups.

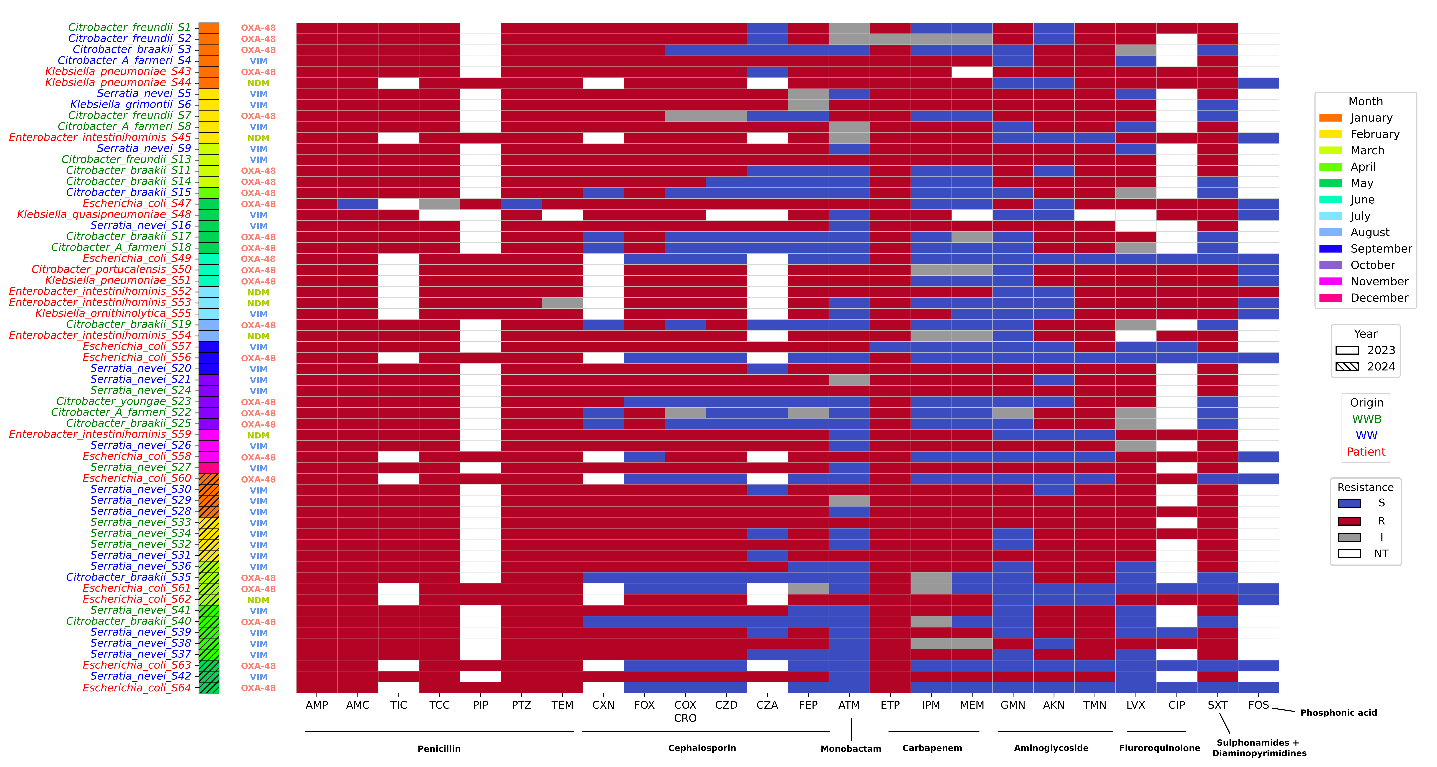

**Supplementary Figure 2: Antimicrobial sensitivity profiles of environmental and clinical isolates**. Heatmap showing susceptibility testing results for environmental (WW and WWB) and clinical isolates according to 2024 CA-SFM/EUCAST clinical breakpoints. Colors represent resistance categories: Resistant (R, red), susceptible with increased exposure (I, grey), Susceptible (S, blue), and Not Tested (NT, white). Rows correspond to individual isolates labeled by Sample ID (Supplementary table 1) and columns represent antibiotics that are grouped according to the antibiotic families. AMP = ampicillin, AMC = amoxicillin-clavulanate, TIC = ticarcillin, TCC = ticarcillin-clavulanate, PIP = piperacillin, PTZ = piperacillin-tazobactam, TEM = temocillin, CXN = cefalexin, FOX = cefoxitin, COX = cefotaxime, CRO = ceftriaxone, CZD = ceftazidime, CZA = ceftazidime-avibactam, FEP = cefepime, ATM = aztreonam, ETP = ertapenem, IPM = imipenem, MEM = meropenem, GMN = gentamicin, AKN = amikacin, TMN = tobramycin, LVX = levofloxacin, CIP = ciprofloxacin, SXT = trimethoprim-sulfamethoxazole, FOS = fosfomycin. The sampling date and sample origin were indicated, with colored boxes and colored text respectively, isolates were represented in chronological order (January 2023 – May 2024).

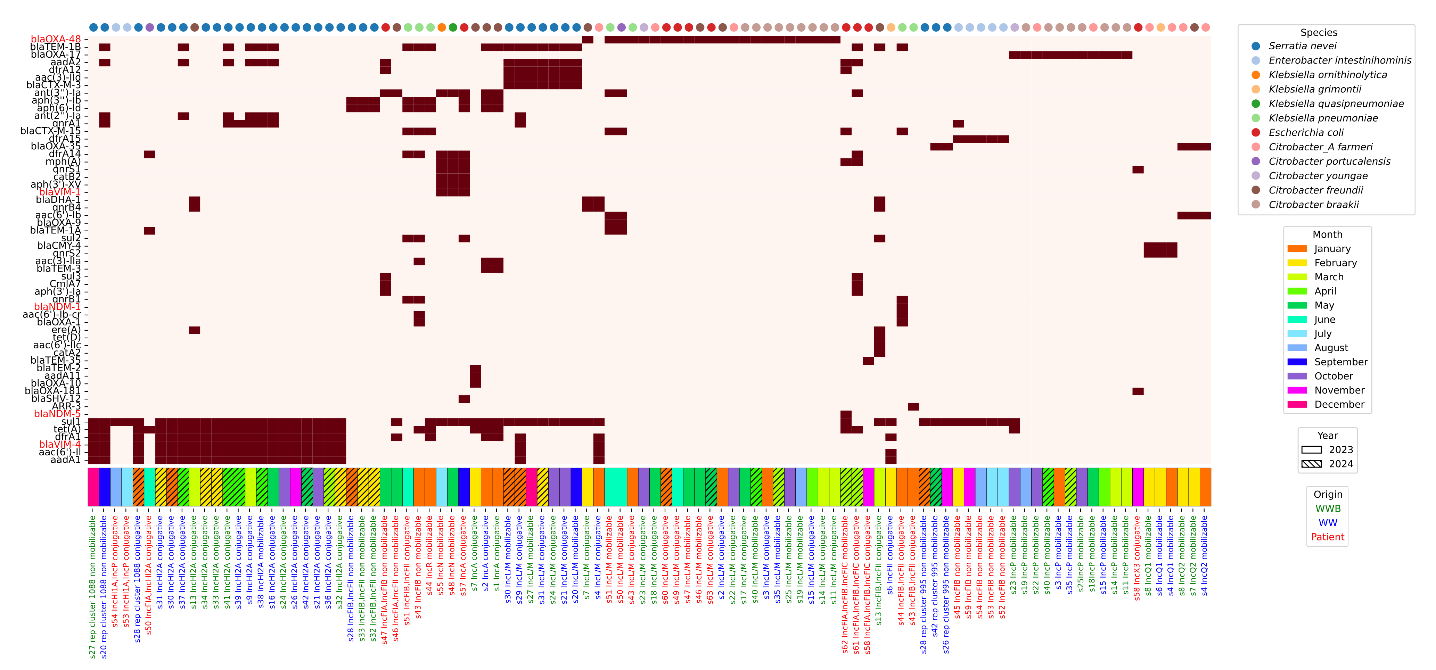

Supplementary Figure 3. Distribution of antimicrobial resistance genes among isolates annotated on plasmids. Resistance genes were identified on plasmid contigs classified by MOB-suite by using the ResFinder database with the abricate software. Carbapenemase genes were annotated in red. Species were attributed with GTDB-Tk classifier and annotated by colored dots for each sample on top of the figure. The sampling date and sample origin were indicated, with colored boxes and colored text respectively. The predicted plasmid type and plasmid mobility were annotated with mobtyper from MOB-suite. The heatmap was obtained with the seaborn module in Python.

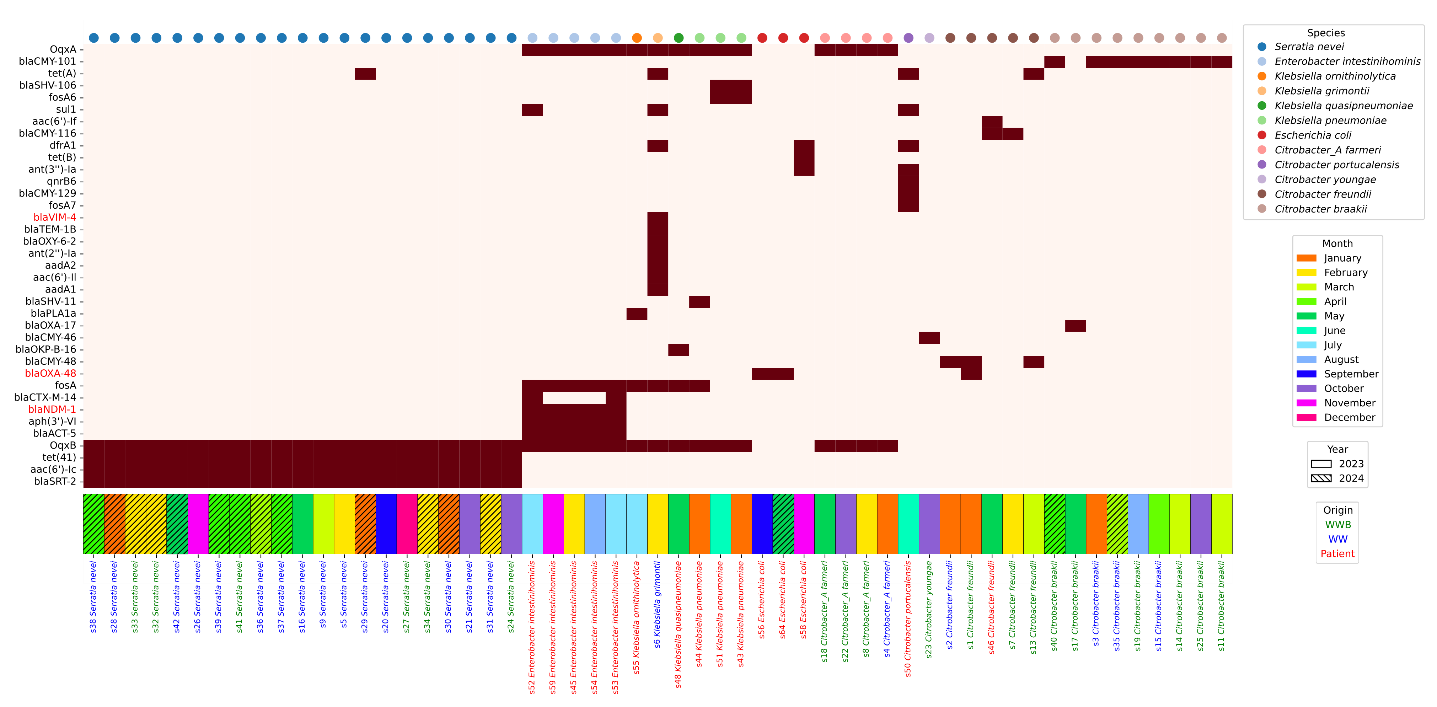

Supplementary Figure 4. Distribution of antimicrobial resistance genes among isolates annotated on chromosomes. Resistance genes were identified on chromosomal contigs classified by MOB-suite by using the ResFinder database with the abricate software. Carbapenemase genes were annotated in red. Species were attributed with GTDB-Tk classifier and annotated by colored dots for each sample on top of the figure. The sampling date and sample origin were indicated, with colored boxes and colored text respectively. The heatmap was obtained with the seaborn module in Python.

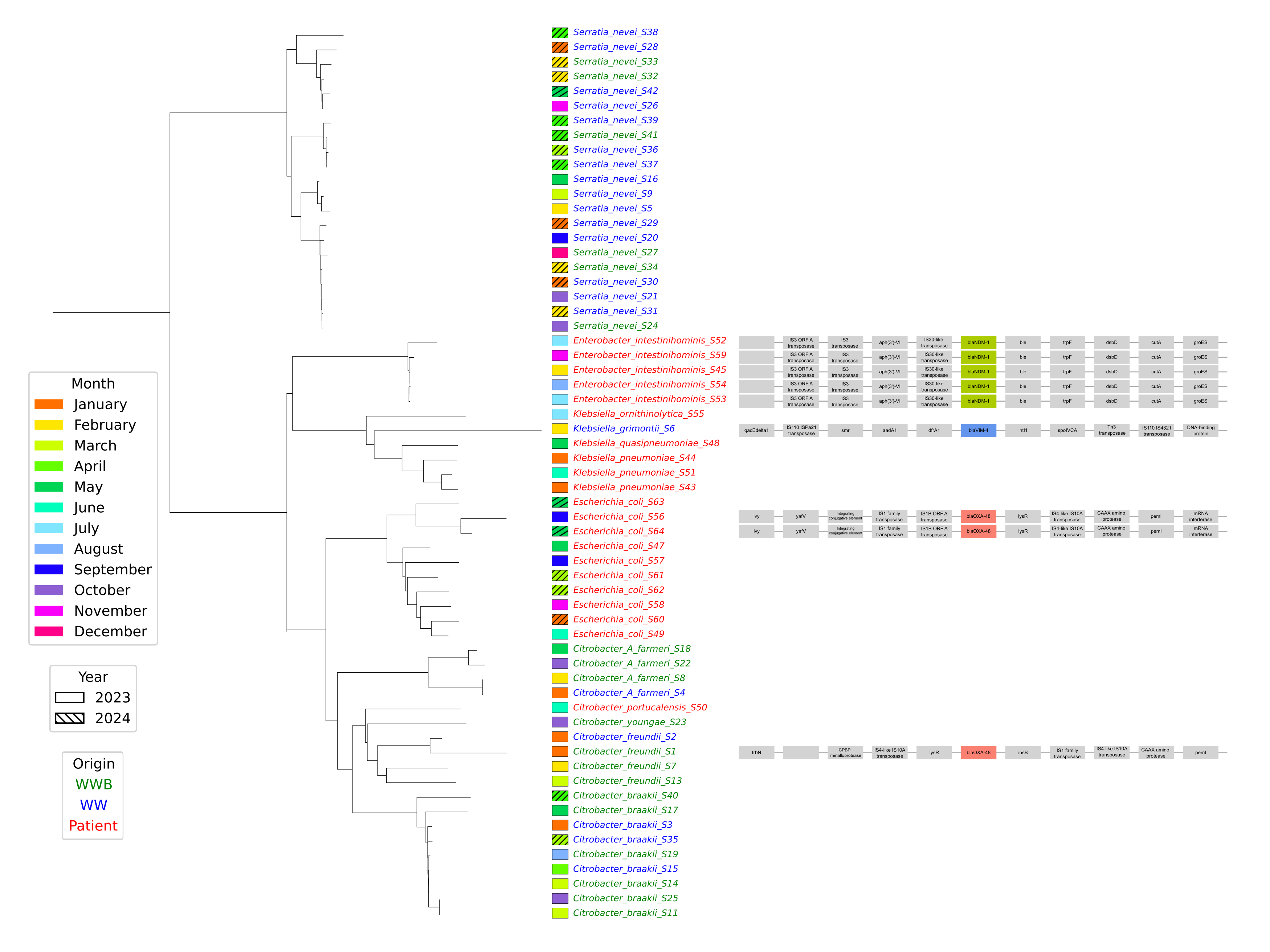

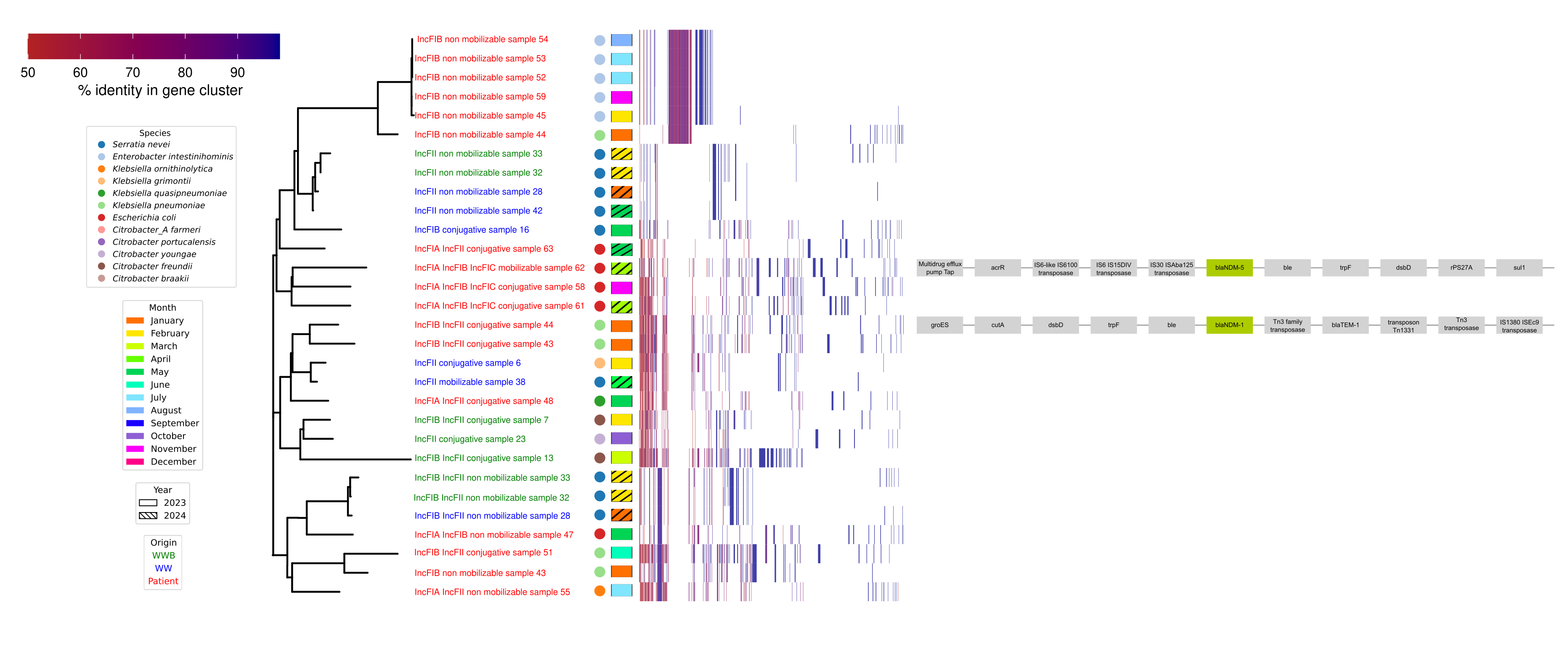

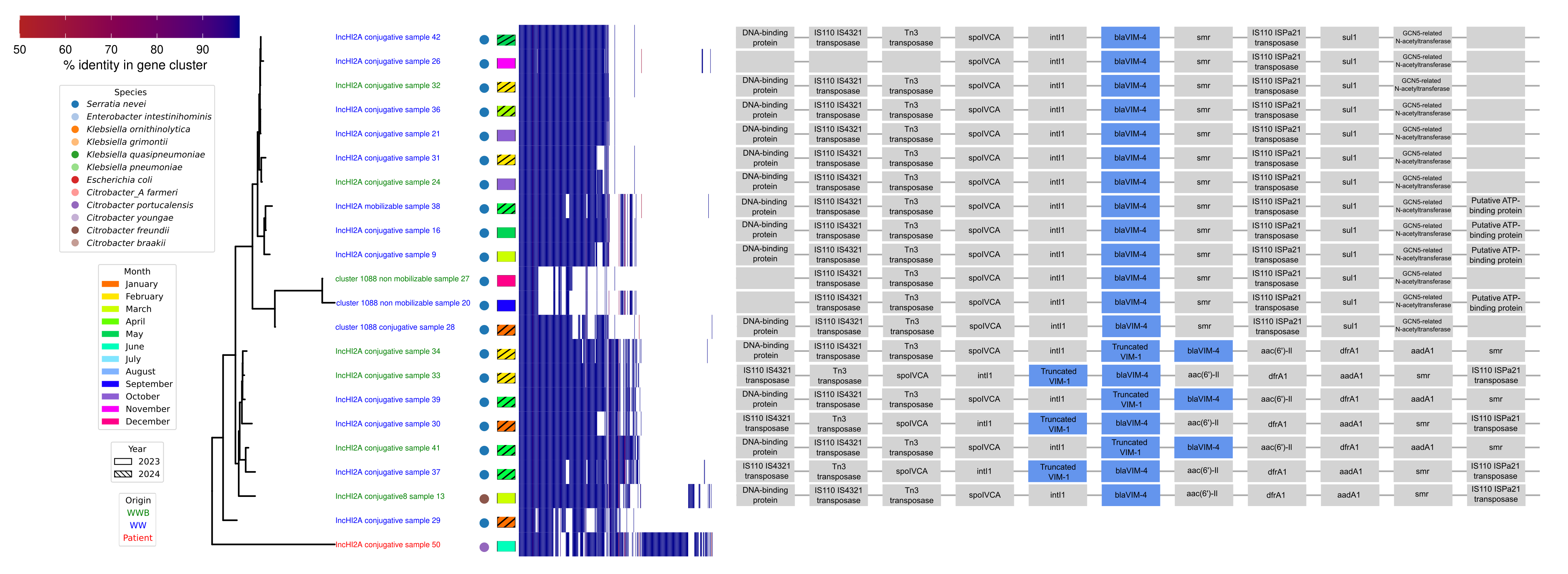

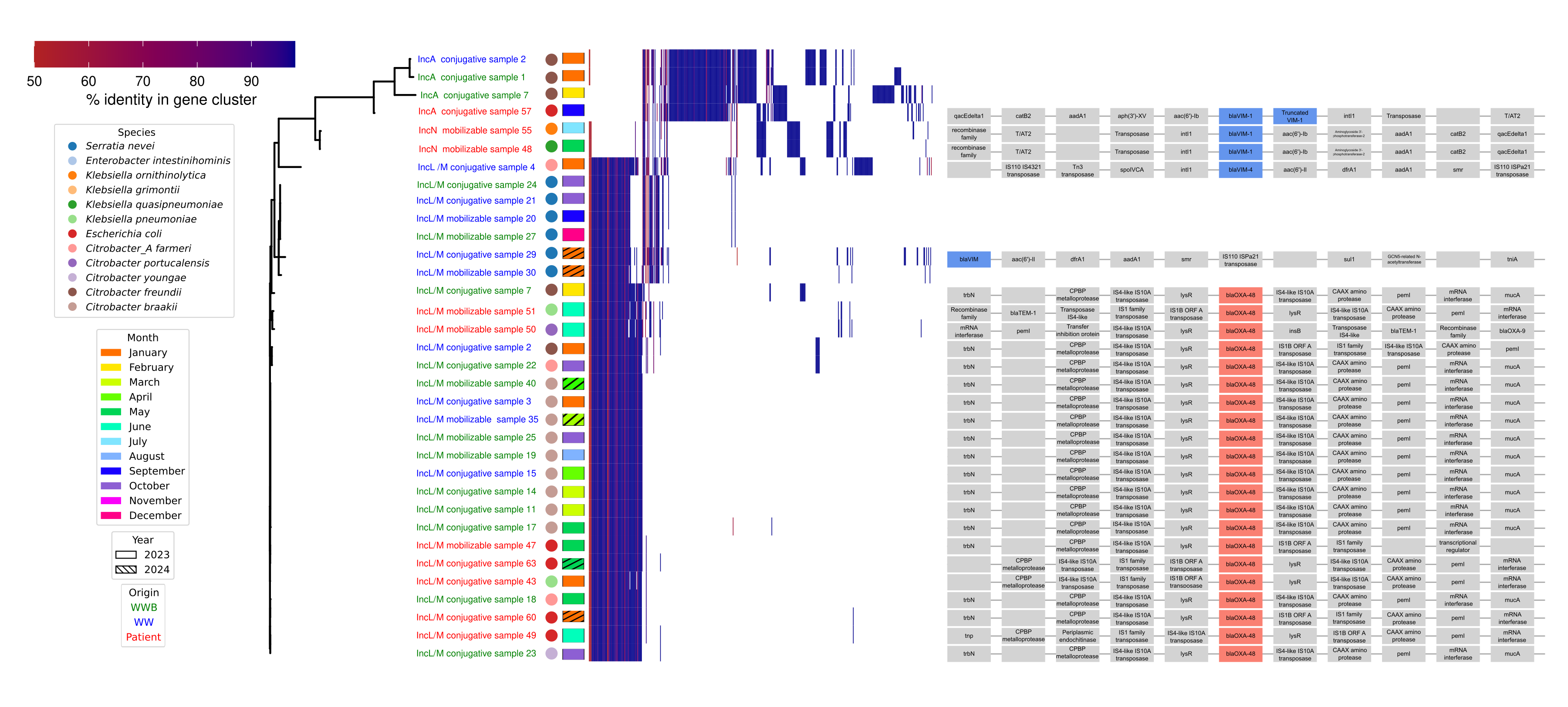

Supplementary Figure 5. Plasmid pangenome-based clustering and genetic context of carbapenemase genes. Pangenome analyses were performed using PIRATE on plasmid contigs identified by MOB-suite and were used to generate a hierarchical clustering tree. The accompanying matrix shows the distribution of gene clusters across plasmids and is colored according to gene-cluster identity. Bacterial species were assigned from chromosomal contigs using GTDB-Tk and are indicated by colored dots. Sampling dates and sample origins are represented by colored boxes and colored isolate labels, respectively. Predicted plasmid types and mobility were determined using mob_typer from MOB-suite. For plasmids carrying carbapenemase genes, the surrounding genetic contexts are displayed alongside the tree. Genes were automatically annotated using Bakta based on the corresponding GFF files. To facilitate comparison among isolates, genetic contexts encoded in the opposite orientation were reverse-complemented and displayed in a consistent left-to-right direction relative to the carbapenemase gene. This graphical reorientation does not indicate a genomic rearrangement. The clustering tree, annotations and genetic contexts were visualized using iTOL.
